# *BRCA1* secondary splice-site mutations drive exon-skipping and PARP inhibitor resistance

**DOI:** 10.1101/2023.03.20.23287465

**Authors:** Ksenija Nesic, John J. Krais, Cassandra J. Vandenberg, Yifan Wang, Pooja Patel, Kathy Q. Cai, Tanya Kwan, Elizabeth Lieschke, Gwo-Yaw Ho, Holly E. Barker, Justin Bedo, Silvia Casadei, Andrew Farrell, Marc Radke, Kristy Shield-Artin, Jocelyn S. Penington, Franziska Geissler, Elizabeth Kyran, Fan Zhang, Alexander Dobrovic, Inger Olesen, Rebecca Kristeleit, Amit Oza, Gayanie Ratnayake, Nadia Traficante, Australian Ovarian Cancer Study, Anna DeFazio, David D. L. Bowtell, Thomas C. Harding, Kevin Lin, Elizabeth M. Swisher, Olga Kondrashova, Clare L. Scott, Neil Johnson, Matthew J. Wakefield

**Affiliations:** The Walter and Eliza Hall Institute of Medical Research, Parkville, VIC, Australia.; Department of Medical Biology, University of Melbourne, Parkville, VIC, Australia.; Fox Chase Cancer Center, PA, USA; Clovis Oncology Inc., San Francisco, CA, USA; School of Clinical Sciences, Monash University, Clayton, Victoria, Australia; University of Washington, Seattle, WA, USA; University of Melbourne Department of Surgery, Austin Health, Heidelberg, Victoria, Australia; The Andrew Love Cancer Centre, Barwon Health, Geelong, Victoria, Australia; Department of Oncology, Guys and St Thomas’ NHS Foundation Trust, London, UK; National Institute for Health Research, University College London Hospitals Clinical Research Facility, London, UK; Princess Margaret Cancer Center, Toronto, ON, Canada; Royal Women’s Hospital, Parkville, VIC, Australia; Sir Peter MacCallum Cancer Centre, Melbourne, VIC, Australia; Sir Peter MacCallum Department of Oncology, University of Melbourne, Parkville, VIC, Australia.; The Daffodil Centre, The University of Sydney, a joint venture with Cancer Council New South Wales, Sydney, New South Wales, Australia; The Westmead Institute for Medical Research, Sydney, New South Wales, Australia; Department of Gynecological Oncology, Westmead Hospital, Western Sydney Local Health District, New South Wales, Australia; QIMR Berghofer Medical Research Institute, Brisbane, QLD, Australia; Department of Obstetrics and Gynecology, University of Melbourne, Parkville, VIC, Australia

## Abstract

*BRCA1* splice isoforms Δ11 and Δ11q can contribute to PARP inhibitor (PARPi) resistance by splicing-out the mutation-containing exon, producing truncated, partially-functional proteins. However, the clinical impact and underlying drivers of *BRCA1* exon skipping remain undetermined.

We analyzed nine ovarian and breast cancer patient derived xenografts (PDX) with *BRCA1* exon 11 frameshift mutations for exon skipping and therapy response, including a matched PDX pair derived from a patient pre- and post-chemotherapy/PARPi. *BRCA1* exon 11 skipping was elevated in PARPi resistant PDX tumors. Two independent PDX models acquired secondary *BRCA1* splice site mutations (SSMs), predicted *in silico* to drive exon skipping. Predictions were confirmed using qRT-PCR, RNA sequencing, western blots and *BRCA1* minigene modelling. SSMs were also enriched in post-PARPi ovarian cancer patient cohorts from the ARIEL2 and ARIEL4 clinical trials.

We demonstrate that SSMs drive *BRCA1* exon 11 skipping and PARPi resistance, and should be clinically monitored, along with frame-restoring secondary mutations.

**Statement of significance:** Few PARPi resistance mechanisms have been confirmed in the clinical setting. While secondary/reversion mutations typically restore a gene’s reading frame, we have identified secondary mutations in patient cohorts that hijack splice sites to enhance mutation-containing exon skipping, resulting in the overexpression of *BRCA1* hypomorphs, which in turn promote PARPi resistance.

## Introduction

Defects in the homologous recombination DNA repair (HRR) pathway, including *BRCA1* and *BRCA2* mutations, are a common feature of high-grade serous ovarian carcinoma (HGSOC) and triple negative breast cancer (TNBC). While *BRCA* defects are known drivers of malignancy in these cancer types, they also make cancer cells susceptible to DNA-damaging platinum agents and poly ADP ribose polymerase (PARP) inhibitor (PARPi) therapy.

PARPi have transformed survival outcomes for many individuals with HRR-deficient (HRD) ovarian cancers; however, drug resistance and disease relapse unfortunately remain common (1–3). The most well-established mechanism of platinum/PARPi resistance for patients with HRD HGSOC is secondary somatic “reversion” mutations in HRR genes that restore the open reading frame disrupted by the primary somatic or germline pathogenic variant. The resulting full length or near full length protein in turn promotes sufficient HRR competency to escape PARPi toxicity (4–9). Other resistance mechanisms have been characterized in preclinical models, with some clinical examples, including expression of hypomorphic *BRCA1* proteins (10,11), loss of the 53BP1-Shieldin axis (12–15), *PARP1* mutations (16), loss of HRR gene methylation (7,17,18), reduced DNA replication gaps (19) and drug efflux (20).

Overall, individuals with *BRCA1*-mutated HGSOC have a worse prognosis than those with *BRCA2*-mutated cancers in the clinic. Among individuals with *BRCA1*-mutated HGSOC, those with frameshift mutations within exon 11 of *BRCA1* have a worse cumulative survival, as well as platinum response, compared with individuals with frameshift mutations outside exon 11 (11,21). This may be explained, in part, by therapy resistance that can arise from the overexpression of *BRCA1* splice isoforms missing most or all of exon 11 (also known as exon 10, but referred to herein as exon 11) (11). The *BRCA1* delta 11q (Δ11q) isoform lacks a large portion (c.788-4096) of exon 11 due to splicing at an alternative donor splice site within the exon, resulting in generation of a shorter but partially functional BRCA1 protein. The delta 11 (Δ11) isoform of *BRCA1* is missing all of exon 11 (c.671-4096), and in human cells is less abundant relative to Δ11q (22–24). However, there is evidence that Δ11 can also partially compensate for loss of full length *BRCA1* (25–27), particularly in a *TP53*-deficient context (27,28).

Canonical *BRCA1* transcripts harboring frameshift exon 11 variants are typically degraded via nonsense-mediated decay (NMD) (29). However, *BRCA1* Δ11 or Δ11q transcripts lack the pathogenic variant-containing exon and are not subject to NMD. Thus, BRCA1-Δ11 or -Δ11q proteins, although truncated, may cause sufficient levels of HRR to induce PARPi and platinum resistance in a tumoral setting (11).

Approximately 30% of pathogenic germline *BRCA1* variants are estimated to occur in exon 11 (30–33). Thus, the cellular mechanisms modulating Δ11 and Δ11q expression are of significance, given *BRCA1* isoforms can promote PARPi and platinum resistance.

Using a cohort of nine HGSOC, TNBC and Ovarian Carcinosarcoma (OCS) PDX models, cell lines, and genomic data from circulating tumor (ctDNA) samples from individuals who took part in the ARIEL2 and ARIEL4 PARPi clinical trials, we investigated factors that determine PARPi and platinum response in cancers with primary exon 11 mutations. This included one matched PDX pair from a woman with HGSOC, before and after multiple lines of treatment, including both chemotherapy and PARPi. We discovered that two of five PARPi-resistant PDX harbored secondary *BRCA1* splice site mutations (SSMs) that were shown to drive alternative *BRCA1* splicing. *BRCA1* SSMs were also enriched in ctDNA from women with HGSOC who had received prior PARPi in clinical trials. Herein, we demonstrate that upregulation of alternative *BRCA1* isoforms, in some cases via secondary splice site mutations, is a mechanism and potential biomarker of PARPi resistance.

## Results

### PDX models with BRCA1 exon 11 mutations have variable PARPi and platinum responses

To study therapeutic responses in the setting of *BRCA1* exon 11 mutations, we assembled panels of OCS, HGSOC and TNBC PDX models in accordance with Institutional Regulatory Board (IRB) approvals at the Walter and Eliza Hall Institute of Medical Research (WEHI) and Fox Chase Cancer Centers (FCCC). A detailed description of PDX, relative to the samples obtained from patients and their clinical course, with alignment of drug response between PDX and outcome for patients, is summarized in Table 1 and Supp. Table 1. All PDX models were analyzed by BROCA sequencing to confirm germline and somatic mutations. All primary *BRCA1* mutations were germline, apart from one that was somatic (HGSOC #049). PDXs were confirmed to be OCS/HGSOC/TNBC by independent histopathological and immunohistochemical (IHC) review (Supplementary Fig. S1; (7,34)).

**Table 1.** PDX and Patient summaries. All PDX and corresponding patient cancers had primary deleterious mutations in exon 11 of *BRCA1* (now known as exon 10) as found by BROCA panel sequencing. Patient platinum response was classified as refractory (progression during treatment or within 4 weeks of last platinum dose), resistant (progression-free interval (PFI) < 6 months), partially-sensitive (PFI = 6-12 months), or sensitive (PFI > 12 months). PDX drug response classifications are outlined in Supplementary table 2. *BRCA1* Δ11 and Δ11q isoform expression classification for PDX are described in Supplementary tables 3 and 4. HGSOC = high grade serous ovarian carcinoma; TNBC = triple negative breast cancer; OCS = ovarian carcinosarcoma; NR = information not relevant for TNBC; NA = data not available; TFI = treatment free interval; DOD = date of death; FU = follow up.

| Patient ID | Cancer type | BRCA1 mutations (transcript NM_007294.4) | Other mutations (BROCA panel) | Lines of therapy received for HGSOC |  |  |  | First platinum m TFI | Primary platinum status | Survival from diagnosis | D11q expression in PDX | D11 expression in PDX | PDX platinum response | PDX PARPi response |
| --- | --- | --- | --- | --- | --- | --- | --- | --- | --- | --- | --- | --- | --- | --- |
|  |  |  |  | Total for patient | Total prior to PDX | Platinum prior to PDX | PARPi prior to PDX? |  |  |  |  |  |  |  |
| #56 | HGSOC | Germline *BRCA1* : c.895_896delGT | *TP53*: c.963del | 3 | 0 | 0 | No | 69 months | Sensitive | 96 months, DOD | Very Low | Very Low | Complete response | Partial Response |
| #56PP | HGSOC | Germline *BRCA1* : c.895_896delGT and somatic *BRCA1*: c.670+281_1101del (20%) | *TP53* :c.963delA; *PTEN*: c.166delT | 8 | 5 | 2 | Yes |  |  |  | Low | Very high | Stable disease | Progressive disease |
| #206 | HGSOC | Germline *BRCA1* : c.3817C>T | *TP53* :c.451C>T; *BLM* rearrangement | 4 | 1 | 1 | No | 17 months | Sensitive | 39 months at last FU | Very Low | Very Low | Complete response | Complete response |
| #124 | TNBC | Germline *BRCA1* : c.1961delA | *TP53* : c.C844T; *RAD51* : c.T659G | 4 | 4 | 1 | No | NR | NR | NA | Low | Very Low | Sensitive | Sensitive |
| #204 | TNBC | Germline *BRCA1* : c.1105_1106insTC | *TP53* : c.C451T; *NBN*: c.2006_2021delinsA | NA | 0 | 0 | No | NR | NR | NA | Very Low | Very Low | Sensitive | Sensitive |
| #032 | HGSOC | Somatic *BRCA1*: c.1251delT | *TP53* : c.97-1_97-6del; *FANCA* : c.3788_3790del | 7 | 6 | 4 | Yes | 11 months | Partially Sensitive | 91 months, DOD | Moderate | Very Low | Stable disease | Progressive disease |
| #049 | HGSOC | Germline *BRCA1* : c.2475delC and somatic *BRCA1* : c.4096+3A>C (33%) | *TP53* : c.428_429insCA | 7 | 4 | 3 | Yes | 5 months | Platinum resistant | 33 months at last FU | Very high | Very Low | Partial response | Stable disease |
| #196 | HGSOC | Germline *BRCA1* : c.1961delA | *TP53*: c.G796A; *ATM*: c.G1595A | 5 | 5 | 3 | Yes | NA | NA | NA | Low | Very Low | Resistant | Resistant |
| #264 | OCS | Germline *BRCA1* : c.2216_2217delAA | *TP53* : c.1006G>T | 5 | 5 | 3 | Yes | 15 months | Sensitive | 93 months, DOD | Very High | Very Low | Progressive disease | Progressive disease |

HGSOC PDX #206 was derived from a patient who had received one prior line of platinum chemotherapy, and the PDX was sensitive to both platinum and PARPi (Figure 1A). For HGSOC #56, we were able to generate a matched PDX pair: one obtained prior to chemotherapy (PDX #56), and one derived after the patient had received five lines of therapy, including having progressed and relapsed on PARPi (PDX #56PP, post-PARPi) (7) (Figure 1B-E). The original chemo-naïve PDX #56 lineage, and a derivative after a single round of platinum *in vivo*, were both PARPi- and platinum-responsive (Figure 1C-D; Supplementary Fig S2). In contrast, PDX #56PP was refractory to both platinum and PARPi, reflecting the patient’s clinical outcomes (Figure 1E).

**Figure 1.**
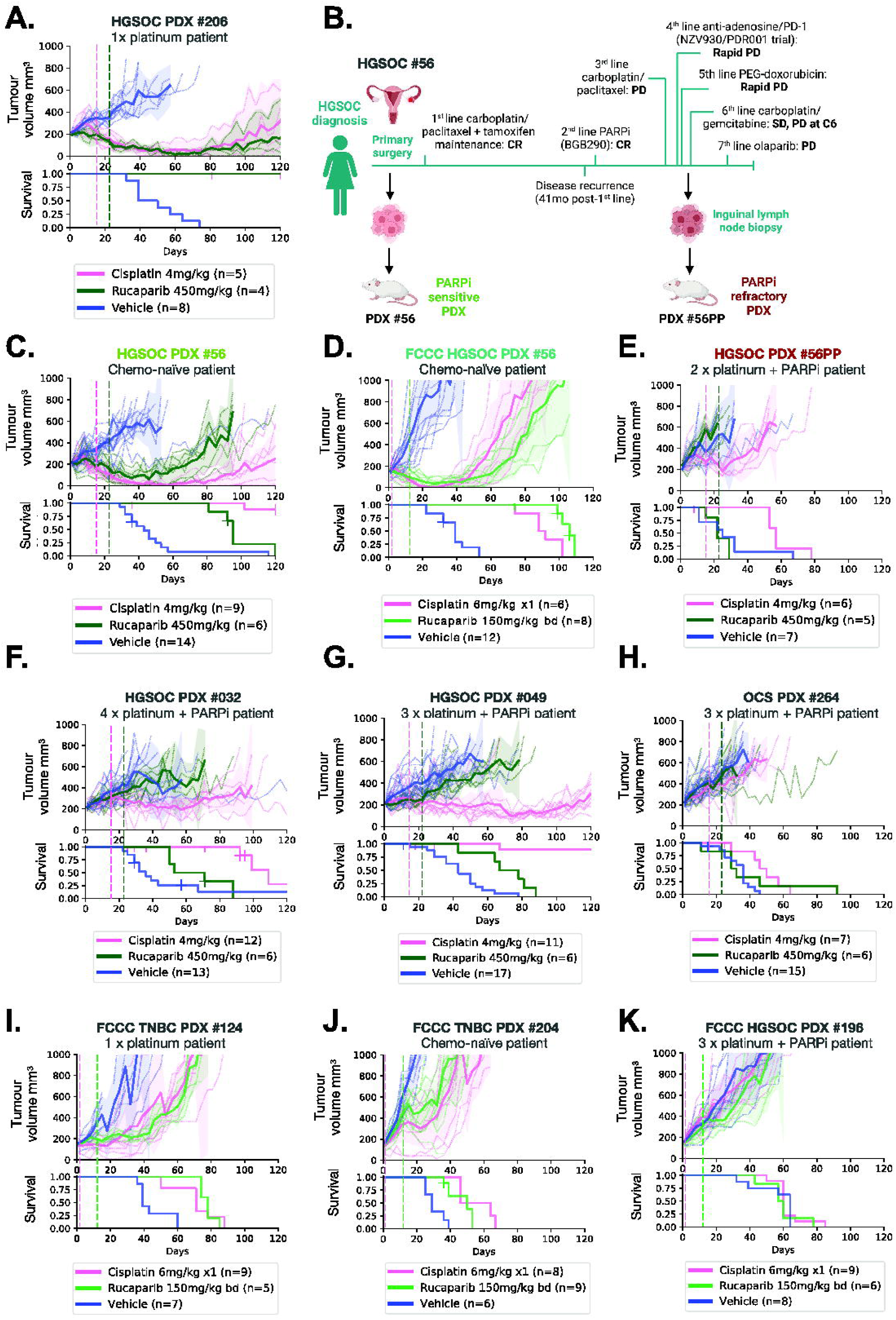
PDX models with *BRCA1* exon 11 mutations from PARPi-treated patients are PARPi-resistant. HGSOC, OCS and TNBC PDX models with *BRCA1* exon 11 mutations were treated with cisplatin or the PARPi rucaparib as indicated by hashed vertical lines, and tumor volume assessed twice weekly. Mean tumor volume (mm^3^) ± 95% CI (hashed lines are individual mice) and corresponding Kaplan–Meier survival analysis. Censored events are represented by crosses on Kaplan–Meier plot; n = individual mice. Detailed patient clinical data can be found in Supplementary table 1. Details of time to harvest, progression and treatment *P* values for each PDX can be found in Supplementary Table 2. **(A)** HGSOC PDX #206 had a complete response to both cisplatin and rucaparib **(B)** Patient #56 timeline, showing generation of the matched HGSOC PDX #56 (chemo-naïve) and #56PP (post-chemotherapy/PARPi patient). Created with BioRender.com. CR = Complete response; PD = Progressive disease; SD = Stable disease; C6 = cycle 6. **(C)** *In vivo* treatment data for HGSOC PDX #56 (*previously published (7)). **(D)** The FCCC derivative of PDX #56 was classified as sensitive to cisplatin and rucaparib, correlating with responses observed for the original lineage treated at WEHI (Figure 1B and Table S2). **(E)** HGSOC PDX #56PP was derived from patient #56 following multiple lines of therapy, including PARP inhibitor, and was refractory to both cisplatin and rucaparib. HGSOC PDX #032 had progressive disease on rucaparib and stable disease on cisplatin. HGSOC PDX #049 had a partial response to cisplatin and stable disease following rucaparib. **(H)** OCS PDX #264 had progressive disease on cisplatin and rucaparib. **(I)** TNBC PDX #124 was classified as sensitive to cisplatin and rucaparib, **(J)** as was TNBC PDX #204. **(K)** HGSOC PDX #196 was resistant to both cisplatin and rucaparib.

HGSOC PDX #032, #049, and OCS PDX #264 were also derived from individuals who were heavily pre-treated, with three-four lines of platinum-based therapy and one line of PARPi prior to PDX establishment, and PDXs were PARPi resistant (Figure 1F-H; Supp Table 1). Interestingly, two of these PDXs still derived some benefit from platinum (HGSOC PDX #049 and #032; Figure 1F-G). HGSOC OCS PDX #264 was unresponsive to both PARPi and platinum (Figure 1H).

FCCC PDX models were treated using different treatment regimens, however similar trends were observed. TNBC PDX #124 was generated from a patient who had received four lines of prior chemotherapy, but only one was platinum-based, and TNBC PDX #204 was from a chemo-naïve patient; both showed a growth delay in response to platinum and PARPi *in vivo* (Figure 1I-J). The OC PDX #196 was derived from a patient who had received 3 prior lines of platinum, and PARPi and showed no response to these drugs *in vivo* (Figure 1K).

In summary, *in vivo* treatment studies of *BRCA1* exon 11 mutated PDX demonstrated a range of PARPi and platinum responses, which correlated largely with prior clinical platinum/PARPi exposure.

### BRCA1 isoform expression analyses in PDX models

We next sought to quantify *BRCA1* exon 11 isoform expression across both cohorts of PDX models. The mRNA expression levels of exon 11-deleted Δ11 and Δ11q isoforms were analyzed by qRT-PCR, as well as total *BRCA1* transcripts measured using primers specific for exon 14 (Figure 2A). Previously described Δ11q-high control cell lines MDA-MB-231, and exon 11 mutated UWB1.289 and COV362 were used as controls (11). PDX #36 is a *BRCA1* WT PDX control.

**Figure 2.**
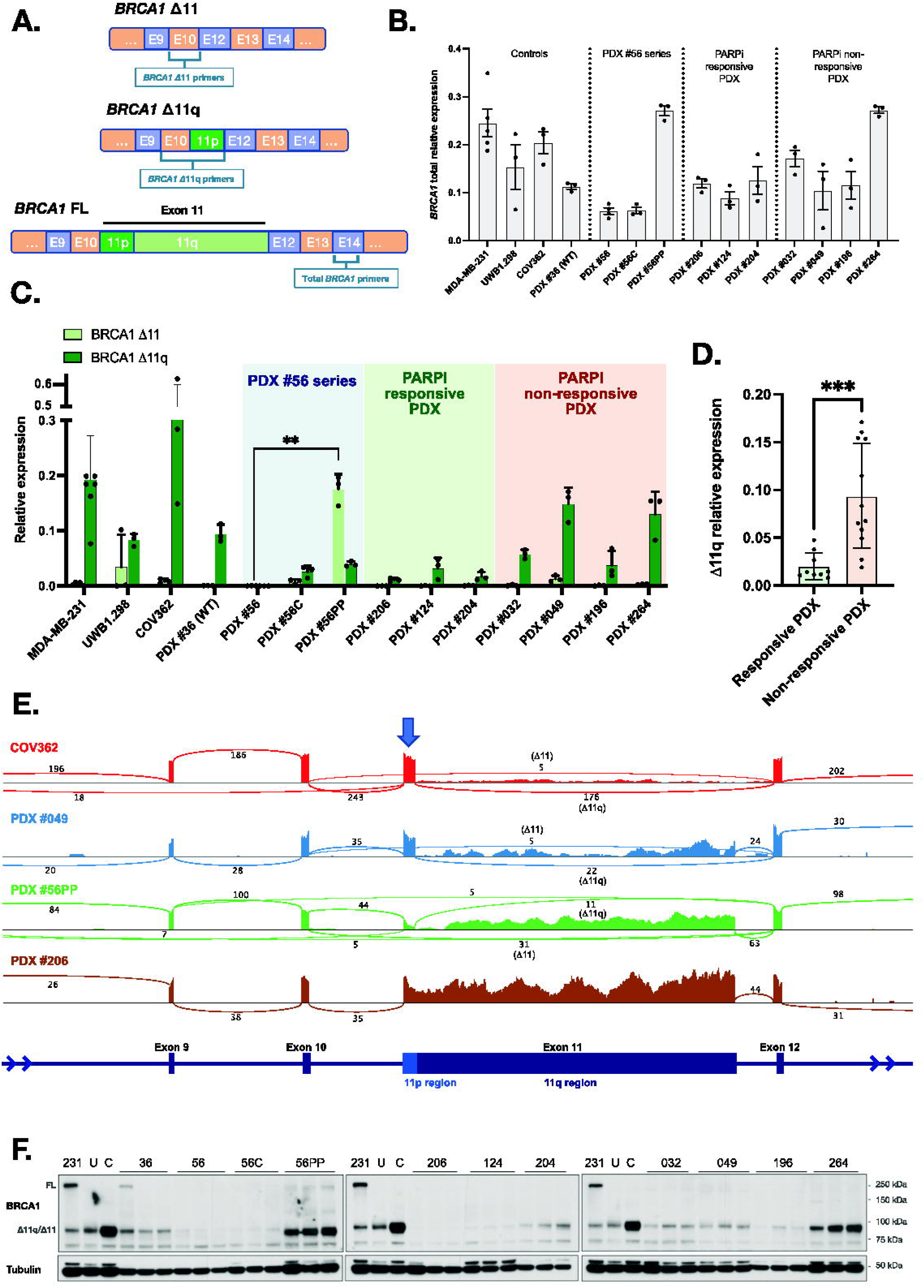
High *BRCA1* Δ11q/Δ11 isoform expression observed in PARPi-resistant PDX models. **(A)** Schematic for design of quantitative two-step PCR (qRT-PCR) assays. **(B)** Relative total *BRCA1* expression for each PDX (mean ± SD). D11q-high cell line MDA-MB-231 (11) was included as a technical control on each qRT-PCR plate, while UWB1.289 and COV362 were included as additional cell line controls. PDX #36 was included as a D11q-high PDX control. **(C)** Relative D11 and D11q expression for each PDX (mean ± SD). PDX #56PP had the highest D11 expression relative to other PDX (*P*=0.0079 compared to matched PDX #56), while PDX #049 and #264 had the highest D11q levels relative to other PDX (classifications in Supp. Tables 3 and 4). **(D)** D11q levels in PARPi responsive PDX models (<2 prior lines of platinum in patient) were lower than levels in non-responsive PDX models (³2 prior lines of platinum + PARPi) (*P*=0.0007). **(E)** RNA sequencing results for a subset of models with either high D11q (cell line COV362, PDX #049), high D11 (PDX #56PP) or low D11/ D11q (#206) and presented as sashimi plots. *BRCA1* exon regions are displayed at bottom of plot, and sequencing read coverage across each exon is represented as a histogram above. The lines connecting exons represent splicing events detected, and the numbers indicate the number of reads assigned to a given event. The blue arrow indicates the exon 11p region, which is retained in the D11q isoform. **(F)** Lysates from nuclear extracts from 3 independent tumors were probed for BRCA1 expression by immunoblotting. Bands at the anticipated sizes for full length (FL) BRCA1 and the D11/D11q isoforms are marked. Tubulin immunoblotting is included as a loading control. Gels were run simultaneously with cell line lysates included as controls for each gel. MDA-MB-231 (231) cells are a BRCA1 wild-type control with full-length (FL) and D11q expression, UWB1.289 (U) and COV362 (C) cells with exon 11 variants and D11/D11q expression. Statistical comparisons were made using an unpaired t-test with Welch’s correction.

Total *BRCA1* expression was variable, with models #56PP and #264 showing the highest levels (Figure 2B). PARPi-resistant HGSOC PDX #049 and #264 tumors were found to express high levels of Δ11q (Figure 2C; Table 1; Supp Table 3-4). PARPi-unresponsive HGSOC PDX models #032 and #196 had moderate and low Δ11q expression, respectively, relative to the control cell lines (Figure 2C; Table 1). Interestingly, #56PP expressed an abundance of the Δ11 isoform (*P*=0.0079 compared to matched PDX #56). When grouped by treatment responses, platinum- and PARPi-responsive PDX (excluding PDX #56 series) demonstrated low levels of Δ11q transcripts relative to resistant PDX tumors (*P*=0.0007; Figure 2C-D; Table 1).

The mRNA expression patterns observed by qRT-PCR correlated with RNA sequencing (RNAseq) data (Figure 2E), and protein expression (Figure 2F), where PARPi-resistant models from patients with prior exposure to PARPi generally had higher levels of Δ11 or Δ11q expression compared to PARPi-sensitive models. Interestingly, mRNA levels did not always correspond with protein expression. For example, PDX #049 and #264 had similar Δ11q mRNA but protein was much more abundant in the latter, indicating post-translational regulation. Nonetheless, in our cohorts, Δ11 or Δ11q isoform expression correlated with PARPi- and, in some cases, platinum-resistance in PDX models derived from patients who had received prior PARPi and multiple lines of platinum therapy.

### Secondary exon 11 splice site mutations drive BRCA1 **Δ**11 *and* **Δ**11q expression

To investigate whether Δ11 and Δ11q expression is a driver of PARPi and platinum resistance in PDX models, we first explored whether other common resistance mechanisms could be detected. We found that reversion mutations that correct the *BRCA1* reading frame and drug efflux by *ABCB1* overexpression were not detected in any PDX models (Supp Table 2; Supp Table 5; Supp Figure S3). WES and BROCA sequencing data also did not detect mutations in known PARPi resistance genes (see Supplementary Information for details).

Interestingly, while no frame-restoring secondary mutations were detected, HGSOC PDX #049 harbored a somatic secondary *BRCA1* splice site mutation (SSM) at the exon 11 donor site (c.4096+3A>C, at 33% frequency), which has been previously shown to drive high Δ11q expression *in vitro* (35) (Figure 3A; Table 1; Supp Table 1 and 5). Of significance, COV362 cells have a functionally similar SSM (c.4096+1G>A at 100% frequency; Figure 3A; Supp Table 1 and 5) previously shown to drive *BRCA1* Δ11q isoform expression *in vitro* (36). These SSM mutations disrupt the 3’ exon 11 donor splice site sequence, which is expected to abrogate exon 11-intron 11 boundary incision. *In silico* predictions indicate that rather than generating intron 11-retaining full-length transcripts, which are likely subject to NMD, the locus switches to generating Δ11q or Δ11 transcripts. The production of the latter isoforms does not require the mutated splice site, with both exon 11 and intron 11 removed, thereby avoiding NMD (Figure 3A). Notably, COV362 cells had the highest Δ11q isoform expression of all models analyzed by qRT-PCR (Figure 2C) and we found COV362 cells to be relatively PARPi resistant (Supp Figure S4). Of note, COV362 has been described as both PARPi or platinum responsive (16,37) or resistant in the literature (38). Our derivative, obtained from ATCC, is PARPi and platinum resistant by CellTiter-Glo viability assays (Supp Figure S4).

**Figure 3.**
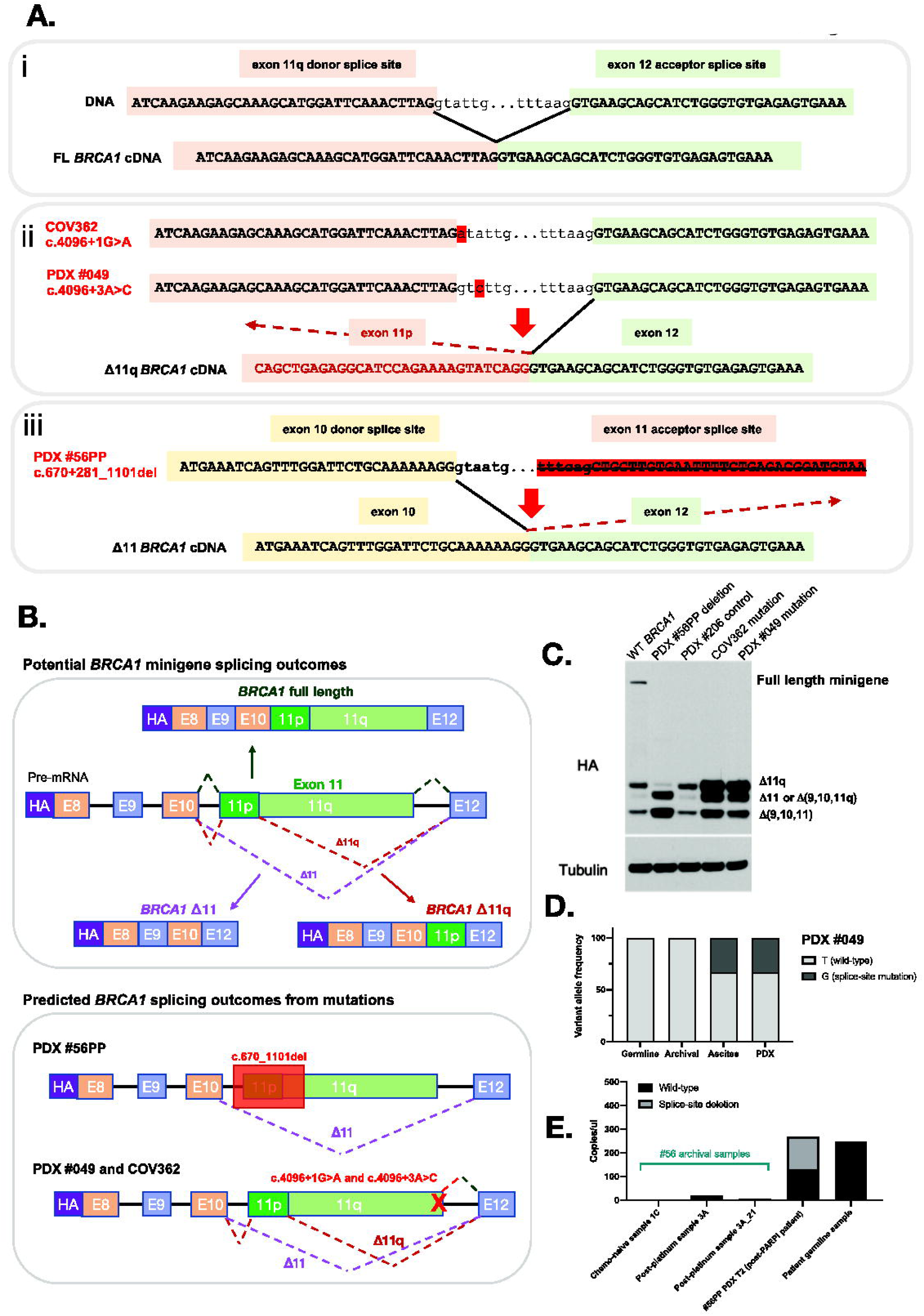
Secondary splice site mutations drive alternative *BRCA1* isoform expression. **(A)** Schematic in section i) shows the *BRCA1* exon 11q donor splice site and its interaction with the exon 12 acceptor splice site under wildtype conditions with no exon skipping. When the donor site is disrupted by a mutation (section ii), like in PDX #049 and COV362, the exon 11p donor site can be used preferentially, thus leading to 11q region skipping. In PDX #56PP the exon 11 acceptor site is disrupted by a large deletion, thus the next acceptor site for exon 12 may be used instead, leading to full exon 11 skipping (section iii). (B) This schematic shows the *BRCA1* mini-gene design, and splicing outcomes predicted for each secondary splice site mutation found in PDX and cell line models. **(C)** Splicing predictions for each secondary splice-site mutation modelled by the mini-gene were confirmed by immunoblotting for the HA tag. The PDX #56PP deletion was confirmed to drive high D11 and potentially also isoform D(9,10,11q) expression. COV362 and PDX #049 secondary splice site mutations were confirmed to drive high D11q relative to the wild-type (WT) *BRCA1* control and the primary deleterious *BRCA1* mutation found in PDX #206. **(D)** Archival material (pre-PARPi) for patient #049 was screened for the secondary splice site mutation found in PDX #049. While the ascites from which the PDX was derived harboured the splice site mutation, the archival sample did not. **(E)** Archival material from the original debulking surgery (1C), or in a sample collected after 1^st^ line platinum therapy (3A and a second DNA extraction 3A_21), for PDX #56PP was tested for the secondary splice site deletion found in the PDX using highly sensitive droplet digital PCR. While no deletion was detected in archival samples, poor droplet amplification in these samples limits interpretation. T2 = transplant/passage 2 PDX aliquot.

In addition to the models harboring exon 11 splice donor site mutations, PARPi-refractory HGSOC PDX #56PP harbored a secondary splice site deletion of 820bp (c.670+281_1101del) crossing the 5’ exon 11 acceptor splice site, which was not present in chemo-naïve and PARPi-responsive PDX #56 (Figure 3A; Supp Table 1 and 5). This mutation was predicted *in silico* to either cause (a) a cryptic acceptor splice site in *BRCA1* exon 11 resulting in a premature stop at codon 234 or (b) skipping of *BRCA1* exon 11 resulting in an in-frame 3426bp deletion and production of the *BRCA1* Δ11 isoform. qRT-PCR results confirmed high expression of the *BRCA1* Δ11 isoform in HGSOC PDX #56PP compared to other models (Figure 2C-E), supporting prediction (b) – exon skipping of the pathogenic germline *BRCA1* mutation.

To test whether the SSMs detected in COV362, HGSOC PDX #049 and HGSOC PDX #56PP were the true drivers of alterative isoform expression in these models, we introduced these mutations into a previously described *BRCA1* minigene system (11) (Figure 3B). In line with *in silico* predictions, SSMs c.4096+1G>A (present in COV362), and c.4096+3A>C (present in PDX #049), led to elevated Δ11q isoform expression relative to controls (Figure 3C), while the c.670+281_1101del deletion (PDX #56PP) led to an increase in Δ11, and Δ9/10/11q (missing exons 9, 10 and 11q region) isoforms (Figure 3C). Indeed, 5 reads of Δ9/10/11q and 31 reads of Δ11 were reported in #56PP by RNAseq (Figure 2D).

Next, we aimed to determine whether these mutations were detectable in archival HGSOC samples from corresponding patients that were collected prior to chemotherapy (indicating whether this mechanism was pre-existing or acquired). Targeted sequencing confirmed the presence of the c.4096+3A>C mutation in the patient ascites sample used to generate PDX #049, but this mutation was not detected in a pre-PARPi, but post-neoadjuvant chemotherapy, archival surgical #049 sample (Figure 3D). This sample was 20% tumor purity (based on TP53 mutation sequencing) and was sequenced to a depth of 68,400x.

The larger secondary splice site deletion in PDX #56PP was measurable by digital droplet PCR and detected with 50% frequency, consistent with a heterozygous state at a *BRCA1* copy number of 2 (Figure 3E; copy number from SNP array data). The matched chemo-naïve PDX #56 did not harbor this deletion, nor did the patient’s archival FFPE chemo-naïve tumor (sample 1C), or a tumor collected five years after 1^st^ line platinum therapy (prior to PARPi) (sample 3A). Whilst the deletion was not detected in either of these pre-PARPi FFPE samples, the DNA samples generated from FFPE appeared to be poor quality, with limited droplet amplification of these samples compared to PDX and germline DNA (Figure 3E). These data suggest that the secondary SSMs in PDX #56PP and #049 were acquired following treatment of patients with chemotherapy/PARPi, although the poor quality of archival FFPE samples limits our assessment. Elevated HRD Sum Scores found in these PDX models (Supp Figure S5), indicating current or historic HRD, may also support an acquired mode of resistance in these initially therapy-responsive patients, although the effects of Δ11 and Δ11q isoforms alone on genomic signatures is not established.

While PARPi-resistant HGSOC PDX #56PP and HGSOC PDX #049 had secondary splice site mutations that explain the high levels of alternative isoform expression, HGSOC PDX #032 and OCS PDX #264 also had high expression of Δ11q, but without driver SSMs in *BRCA1*. Certain single nucleotide polymorphisms (SNPs) have been reported to cause elevated expression of the *BRCA1* Δ11q isoform (23,39,40), and we investigated these in whole exome sequencing (WES) data for the other PARPi-resistant models HGSOC PDX #032 and OCS PDX #264. None of the SNPs/variants investigated from the literature were identified in these PDX, suggesting that additional mechanisms exist for elevating *BRCA1* Δ11q expression.

### BRCA1 *Δ*11 *and Δ*11q isoform expression drives PARPi resistance

The BRCA1-Δ11q protein is hypomorphic and its expression in BRCA1 deficient cells does not restore HR to the same extent as expression of full-length BRCA1 (11). Therefore, we aimed to measure the effects of BRCA1-Δ11q expression in our cell line and PDX models on therapy sensitivity. To do this, we first utilized targeted silencing of the *BRCA1* Δ11q isoform using siRNA followed by PARPi treatment in COV362 (Figure 4A). A clear reduction in colony forming capacity was observed in COV362 cells treated with a broad *BRCA1* siRNA and targeted Δ11q siRNA compared to the scrambled control cells (Figure 4B-D; Supp Figure S6-7). Silencing of *BRCA1* and the Δ11q isoform was confirmed using qRT-PCR (Supp Figure S6). These results reveal that high *BRCA1* Δ11q isoform expression, driven by a secondary SSM, contributes to PARPi resistance. We also showed that COV362 formed RAD51 foci following DNA damage, a biomarker of HR DNA repair and PARPi resistance (Figure 3E-G; Supp Figure S8-9).

**Figure 4.**
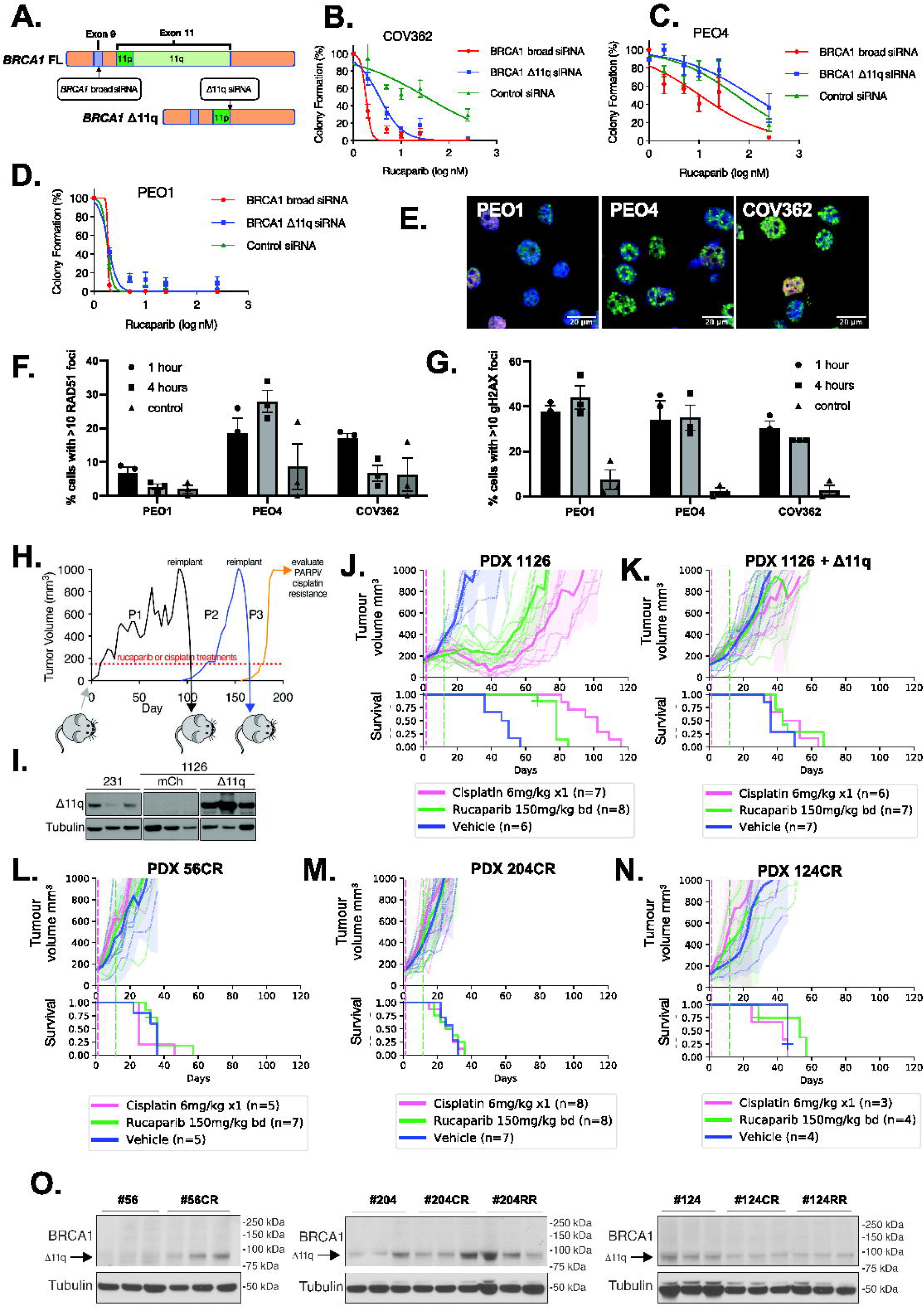
*BRCA1* Δ11q isoform expression drives PARPi resistance in cell line and PDX. **(A-D)** COV362 was sensitised to PARPi rucaparib following siRNA knockdown of either D11q specifically, or broad knockdown *BRCA1*, with schematic of siRNA design shown in (A). Mean ± SEM are shown for three independent colony formation experiments. Knockdown was confirmed by qRT-PCR (Supp Figure S6). Controls are PEO1 (HR deficient) and PEO4 (HR proficient). **(E-F)** COV362 demonstrated some capacity to form RAD51 foci relative to control 1 hour after DNA damage (10 Gy irradiation; not significant; *P* value = 0.153). **(G)** DNA damage following irradiation was confirmed by gH2AX foci (mean ± SEM percent of geminin positive cells with ≥10 RAD51 or gH2AX foci). **(H)** PDX #1126 (BRCA1 exon 13 mutation) tumors expressing either mCherry or BRCA1-D11q were treated with cisplatin or rucaparib. Treated tumors were transplanted into recipient mice for further treatment (Supp Figure S10B-C). **(I)** BRCA1-D11q protein expression was assessed in PDX #1126 infected with BRCA1-D11q or mCherry encoded lentivirus following *in vivo* selection with cisplatin. MDA-MB-231 (231) lysate was included as a control. **(J)** Cisplatin, rucaparib and vehicle responses were assessed in PDX #1126. Treatment end = vertical hashed lines. Mean tumor volume (mm^3^) ± 95% CI (hashed lines are individual mice) and corresponding Kaplan–Meier survival analysis. Censored events = crosses on Kaplan–Meier plot; n = individual mice. **(K)** Cisplatin and rucaparib responses for PDX #1126 tumors with ectopic BRCA1-D11q expression following *in vivo* selection for cisplatin resistance. **(L)** PDX #56 was subjected to *in vivo* selection for cisplatin resistance as described in (H). Cisplatin and rucaparib responses following 3 passages of cisplatin re-treatment of PDX #56 (PDX #56CR). **(M)** Cisplatin and rucaparib resistant PDX #204 (PDX #204CR) were obtained following 3 passages of cisplatin re-treatments of PDX #204. Resistance was similarly obtained using rucaparib re-treatments (Supp Figure S10E). **(N)** Cisplatin resistant PDX #124 (PDX #124CR) was obtained following 4 passages of cisplatin re-treatment. Resistance via rucaparib re-treatments was also achieved (Supp Figure S10F). **(O)** Three independent tumors per PDX lineage were probed for BRCA1 expression by immunoblotting. Bands at the anticipated sizes for full length (FL) BRCA1 and the D11/D11q isoforms are marked. Tubulin immunoblotting is included as a loading control. Corresponding mRNA analysis is presented in Supp Figure S10E.

To determine whether the Δ11q isoform drives PARPi resistance *in vivo* and in the PDX setting, we induced ectopic Δ11q or mCherry control expression in the BRCA1-null PARPi/platinum-sensitive TNBC PDX #1126, which has a primary exon 13 frameshift mutation (41) (Figure 4H-K; Supp Table 2). Following Δ11q or mCherry infection, serial cisplatin treatments were used to drive resistance in the lineage with added Δ11q (Figure 4H-I, Supp Figure S10). The same approach did not result in resistance in the lineage with mCherry. The post-platinum treated Δ11q-expressing PDX expressed abundant ectopic Δ11q and lacked response to both PARPi and platinum compared to the isogenic mCherry-expressing PDX (Figure 4K).

Moreover, we applied serial cisplatin or PARPi treatments to HGSOC #56, TNBC #204 and TNBC #124 and developed derivatives that were PARPi (RR) and cisplatin resistant (CR) (Figure 4L-N, Supp Figure S10). Elevated Δ11q expression was observed in some resistant tumors derived from PDX #56 and #204, however no Δ11q increase was observed in resistant PDX #124 (Figure 4O; Supp Figure S10E). Of note, #56CR, unlike PDX #56PP had elevated Δ11q without a detectable secondary mutation, suggesting multiple mechanisms for elevating Δ11q can evolve from the same tumor.

In summary, we confirmed that exon-skipping alternative *BRCA1* isoforms are drivers of PARPi resistance in our models, and this mechanism could be acquired *in vivo* following platinum/PARPi treatment.

### Secondary BRCA1 SSMs enriched post-PARPi in tumors and ctDNA

To explore the potential clinical relevance of SSMs, we examined tumor and ctDNA samples from patients enrolled in the clinical trials ARIEL2 (NCT01891344; individuals with platinum-sensitive relapsed HGSOC, Fallopian Tube, or Primary Peritoneal cancer) and ARIEL4 (NCT02855944; individuals with *BRCA1/2*-mutated relapsed ovarian cancer) before and after treatment with the PARPi rucaparib.

Of the patients from the ARIEL2 and ARIEL4 trials with *BRCA1* exon 11 mutated cancer, there were 115 individuals for whom pre-PARPi (rucaparib) tumor/ctDNA samples with sequencing data were available, and 63 matched patients for whom post-PARPi data were also available (Figure 5A). Of the pre-PARPi (but post-platinum) samples, 8% (9/115) had a frame-correcting secondary *BRCA1* mutation, and 1% (1/115) had an SSM impacting the *BRCA1* exon 11 donor splice site (c.4096 region). Following PARPi, this increased to 29% (18/63) with frame-correcting secondary mutations, and 8% (5/63) with SSMs (Figure 5A). *In silico* modelling of SSMs predicted that all would drive expression of the Δ11q isoform (Figure 5B-C; Supp Table 5), and this was confirmed using the *BRCA1* minigene system (Figure 5D).

**Figure 5.**
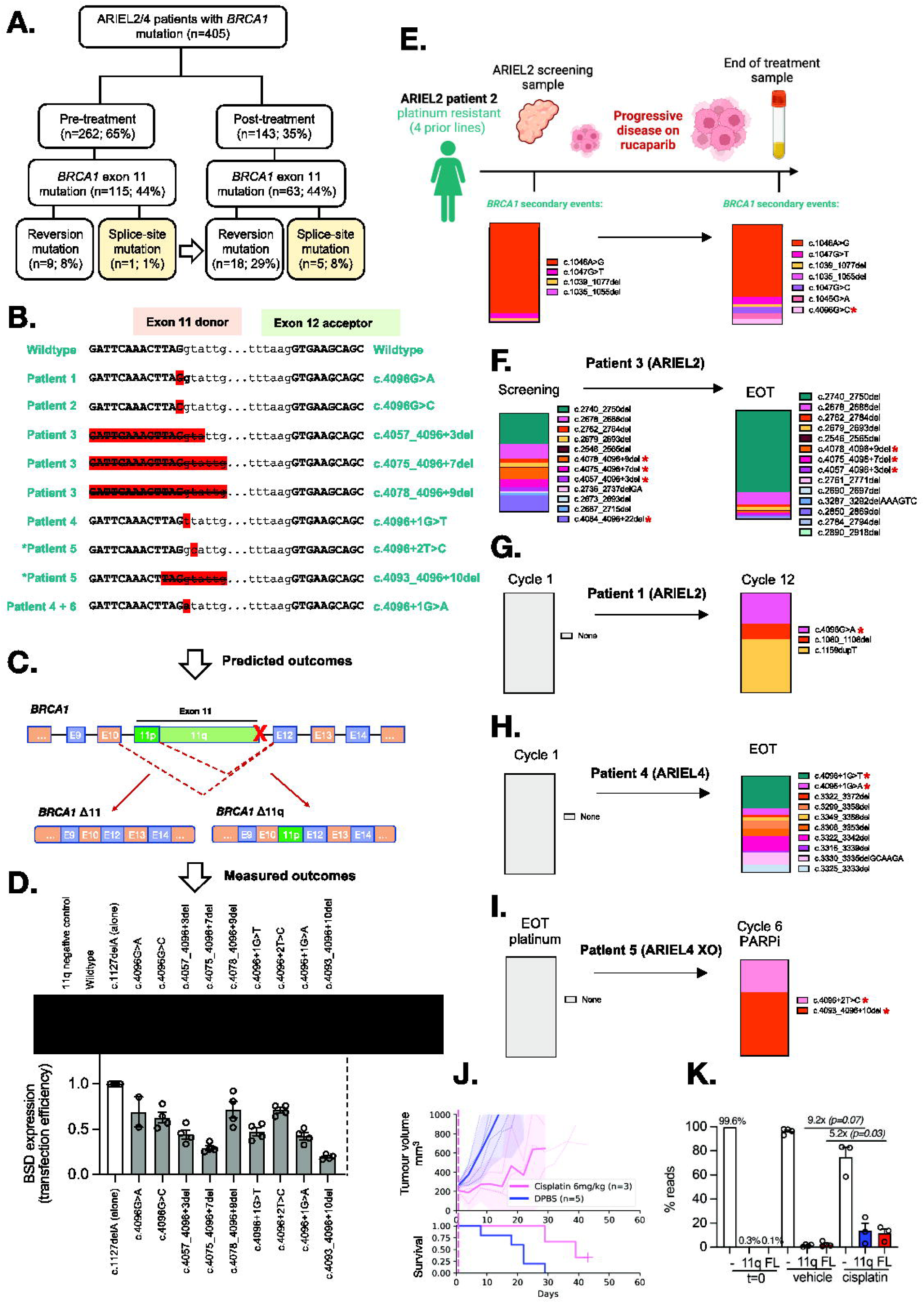
*BRCA1* secondary splice-site mutations are enriched in ARIEL2/4 clinical trial patient samples following PARPi treatment. **(A)** *BRCA1* secondary splice-site mutations increased form 1% (pre-PARPi) to 8% (post-PARPi) in patient tumor/plasma samples from the ARIEL2 and ARIEL4 clinical trials. **(B)** The *BRCA1* (NM_007294.4) exon 11 donor splice-site mutations identified in these patients and the DNA sequence context are presented. *Patient 5 is an ARIEL4 chemotherapy to PARPi cross-over (XO) arm patient, not included in part A (n=5). **(C-D)** The predicted outcomes based on these disruptions of the exon 11 donor splice site (detailed in Supp Table 9) were confirmed for most mutations (D) using the previously described *BRCA1* minigene system (11). Mutations driving lower levels of D11/D11q also had a reduced minigene transfection efficiencies relative to other samples. **(E)** Summary of *BRCA1* secondary events detected in Patient 2 (ARIEL2) before and after PARPi therapy, and their relative proportions in each sample (by colour). **(F)** A number of *BRCA1* secondary events were also detected in the screening biopsy for platinum and rucaparib resistant Patient 3 (ARIEL2) prior to PARPi, and the number increased at end of rucaparib treatment (EOT). **(G)** In contrast, platinum resistant (4 prior lines of platinum) Patient 1 (ARIEL2) had no secondary *BRCA1* events detected at first cycle of rucaparib, and had stable disease (SD) on treatment. Three secondary events were detected at cycle 12 of treatment, including a splice-site mutation c.4096G>A. **(H)** Patient 4 (ARIEL4) was partially platinum sensitive (2 prior lines) with no secondary *BRCA1* events detected at cycle 1 of rucaparib. The EOT plasma sample was positive for multiple reversion events and two splice site mutations (4096+1G>T and 4096+1G>A) confirmed by minigene to alter splicing (D). **(I)** Patient 5 (ARIEL4) was platinum resistant and was enrolled in the chemotherapy arm of ARIEL4. They then crossed over to receive rucaparib treatment where they had stable disease. There were no secondary events detected prior to starting rucaparib, but at cycle 6 there were two *BRCA1* splice-site mutations detected (c.4096+2T>C and c.4093_4096+10del), without other reversion events. c.4096+2T>C was found to drive alternative *BRCA1* splicing (D). Red stars (*) indicate SSM’s detected in patient examples (E-I). **(J)** SUM149 cells expressing BRCA1-D11q (11q) or full length BRCA1 (FL) were mixed equally (0.1%) with a BRCA1-null (-) SUM149 derivative cell line (99.8%) (11). These cell mixtures were injected into mice and treated with vehicle (DPBS) or 6mg/kg cisplatin. Mean tumor volume (mm^3^) ± 95% CI (hashed lines are individual mice) and corresponding Kaplan– Meier survival analysis. Censored events = crosses on Kaplan–Meier plot; n = individual mice. **(K)** Targeted sequencing was used to measure the representation of each SUM149 derivative in tumors from (J) and from the cell mixture prior to implantation (t= 0). 11q representation increased 9.2-fold (*P* = 0.07) while FL representation increased 5.2-fold (*P*= 0.03) in cisplatin treated tumors compared to vehicle treated tumors. Each datapoint indicates read percentage for independent tumors and *P* value shown from unpaired t-tests for the indicated comparisons.

Interestingly, in several cases, samples with c.4096 SSMs also harbored multiple additional reversion events following PARPi therapy (Figure 5E-I; Supp Table 8). Unlike sequencing of tissue samples (e.g. ARIEL2 patient 6; Supp Figure S11A), liquid biopsies are not spatially restricted and can sample secondary events across a woman’s entire cancer. Examples of other cases with multiple reversion events, excluding SSMs, are presented in Supp Figure S11.

Since SSM’s and reversion mutations were found in the same samples for some patients, we wanted to examine if either the Δ11q or full length BRCA1 proteins provided a selective advantage, so we performed an *in vivo* competition experiment. We tested the concept that cells containing either a frame-reverting mutation or a splice site secondary mutation might both be subject to positive selection during therapy. SUM149 is a TNBC cell line with a *BRCA1* exon 11 frameshift variant (c.2288delT) that shows high levels of Δ11q expression. We previously made derivatives where Δ11q was knocked-out (null) or a frame-correcting reversion mutation was introduced (11). In the current study, we mixed equal amounts (0.1% each) of Δ11q-high parent cells and “FL” (full length *BRCA1*) cells harboring a secondary mutation, with *BRCA1*-null (-) SUM149 cells (99.8%). These cell mixtures were then injected into mice and treated with vehicle or 6 mg/kg cisplatin (Figure 5J). Sequencing revealed that cells with secondary mutations (FL) or only Δ11q expression (11q) were selected for equally with platinum pressure (Figure 5K), providing a potential rationale for why both reversion and SSM were detected in the same therapy-resistant patient samples.

In addition to SSMs affecting *BRCA1* exon 11, SSMs affecting a different exon, exon 20, were also detected in four individuals enrolled on the ARIEL4 trial. All four cases were found to have the same primary germline *BRCA1* exon 20 pathogenic variant: c.5266dupC (Q1756fs; Supp Table 8). Interestingly, three of the four individuals had the same SSM, *BRCA1* c.5277+1G>T, accompanied by a nearby c.5276A>G variant. The additional c.5276A>G variant may be required to re-establish the boundary sequence adjacent to the splice site to enable proper splicing (Supp Figure S11D).

We observed one additional case with a *BRCA1* SSM predicted to drive skipping of a pathogenic variant in exon 2, and another case with a pathogenic *BRCA2* variant in exon 8 and an SSM predicted to drive skipping of exon 8 (Supp Table 8-9). These additional cases involving other exons of *BRCA1,* and even *BRCA2,* suggest that this same resistance mechanism may be relevant beyond exon 11 of *BRCA1*.

## Discussion

In this study we demonstrated that high *BRCA1* Δ11/Δ11q isoform expression is associated with resistance to PARPi in -HGSOC, OCS, and TNBC. *BRCA1* Δ11/Δ11q expression was also subject to positive selection in PDX models treated with platinum. In some cases, this high expression of alternative *BRCA1* isoforms was driven by SSMs. This includes HGSOC PDX #049 with a secondary donor splice site mutation and very high Δ11q expression, and HGSOC PDX #56PP with a heterozygous secondary deletion across the exon 11 acceptor splice site, driving extremely high Δ11 expression relative to other models.

It is known that certain *BRCA1* splice site variants can drive elevated Δ11 and Δ11q expression *in vitro* (35,36), for example c.4096+1G>A (also called IVS11+1G>A) and c.4096+3A>G (IVS11+3A>G). While these splice site variants have been identified in the germline of multiple families with history of breast and ovarian cancer, evidence of their pathogenicity and impact on inherited cancer susceptibility has been mixed (42,43). Indeed, a healthy homozygous carrier of the c.4096+3A>G variant has been reported (44). Interestingly, a c.4096+1G>A SSM was previously reported in a patient with a primary germline exon 11 mutation (*BRCA1*:c.2043dup) following progression on PARPi therapy (ARIEL3 trial) (45), suggesting that that this SSM may have been driving PARPi resistance.

The PARPi-resistant COV362 cell line was also found to harbor an SSM in *BRCA1* in addition to its primary *BRCA1* mutation, and had extremely high Δ11q isoform expression. COV362 was derived from a pleural effusion of a patient with ovarian endometrioid carcinoma (46), although molecular profiling studies support COV362 falling within the HGSOC subtype (47,48). It is unclear from the original publication whether this patient had received prior chemotherapy (46), and whether the SSM was present in the original carcinoma sample, or acquired through cell culture. Nevertheless, silencing of the Δ11q isoform in the COV362 cell line led to PARPi sensitization, confirming that overexpression of this *BRCA1* isoform is indeed a key driver of PARPi resistance in this model.

Strikingly, SSMs, including the one found in COV362, were found to be enriched post-PARPi exposure in tumor and ctDNA from patients in the ARIEL2/4 clinical trials, providing further clinical evidence that these play a role in PARPi-resistance. In addition to SSMs that cause exon 11 skipping, in other cases we found SSMs in ctDNA predicted to drive *BRCA1* exon 19 skipping and an SSM previously demonstrated *in vitro* to remove exon 7 and 8 of *BRCA2* (49), in each case to remove a pathogenic germline variant from the transcripts. This suggests that the phenomenon is not restricted to exon 11 of *BRCA1*, but may have relevance for other *BRCA1* exons and even other HRR pathway genes.

There were several cases with multiple additional distinct *BRCA1* secondary/reversion events in ctDNA (patients 2, 3 and 4 in Figure 5). HGSOC is known to be a highly heterogeneous cancer type driven by genomic instability which can be exacerbated by therapy. This was reflected clearly in the sheer magnitude of secondary events found in several post-PARPi ARIEL2/4 liquid biopsies. This was indeed a sobering finding, further highlighting the need for development of more therapeutic options for patients with this heterogeneous and highly adaptable cancer type.

Our identification of genomic alterations associated with *BRCA1* alternative isoform expression is particularly important for this PARPi resistance mechanism, as measuring isoform expression in archival/biopsy tumor samples can be difficult and confounded by contamination with normal tissue. Our cell line and PDX models made it possible to compare relative isoform expression using human-specific primers, as the tumors are relatively pure, lacking a non-neoplastic human tissue component. The SSMs identified in our study now make it possible to readily screen for this form of PARPi and platinum resistance in patients by sequencing tissue or ctDNA from liquid biopsies, as we have demonstrated here.

Several PARPi-resistant PDX tumors showed high expression of BRCA1 Δ11q, but lacked SSMs – for example OCS PDX #264 and HGSOC PDX #032. We could not find SNPs known to drive alternative isoform expression in these models, so it is possible that trans-elements, such as splicing factors and regulatory proteins, may be playing a role. We plan to explore these cases in future studies, where we hope to uncover additional clinically useful biomarkers for this form of PARPi resistance. Interestingly, homozygous *Brca1^Δ11/Δ11^* mice are embryonic lethal, and derived cells are PARPi sensitive (25–27). Therefore, it is likely that additional re-wiring events accompany SSMs, such as reduced expression of end resection inhibitory proteins, which work in conjunction with the BRCA1 Δ11 or Δ11q protein to promote robust HR and PARPi resistance (50,51). Alternatively, in the setting of cancer, BRCA1 Δ11 or Δ11q expression and the residual HR it provides is sufficient to induce PARPi resistance in tumors.

The utility of spliceosome inhibitors combined with PARPi should be explored in cancers where alterative *BRCA1* isoforms are identified as a driver of drug resistance. Pladienolide-B has been previously shown to reduce Δ11q expression *in vitro* (11). However, the classical small molecule inhibitors of the splicing machinery (e.g. pladienolide-B, spliceostatin A, GEX1A and E1707) have not been clinically useful, due to their high toxicity. Novel spliceosome inhibitors with reduced toxicity profiles, such as the SF3B1-modulator H3B-8800, may provide some hope for patients with Δ11q-driven PARPi resistance (52–54).

Our data clarifies the importance of transcriptional splicing plasticity, as a cause of drug resistance and illustrates significant challenges for the genomic medicine field. Clinical genomics reports usually only refer to one canonical transcript per gene, masking significant complexity with clinical impact. The functional consequence of a wider array of mutations requires real-time resolution and reporting, for accurate clinical guidance. The Multiplex assays of variant effect (Mave) database (MaveDB; https://www.mavedb.org), a public repository for large-scale measurements of sequence variant impact, may assist (55).

In conclusion, alternative *BRCA1* isoform expression is a driver of PARPi resistance across multiple cancer types. Our preclinical experiments and analysis of samples from the ARIEL2/4 clinical trials also establish the role of *BRCA1* secondary splice site mutations in driving this form of PARPi resistance. These SSMs should be screened for in people with PARPi-resistant cancers, so that if found, alternative treatment options may be explored. Pre-emptive therapies to avert this cause of PARPi resistance are also urgently needed.

## Methods

### Reagents

Rucaparib was kindly provided by Clovis Oncology. The following antibodies were used for WEHI PDX immunohistochemistry: p53 (M700101 1:100; Dako), Ki67 (M7240 1:50; Dako), Cytokeratin (Pan-CK; M3515 1:200; Dako), PAX8 (10336–1-AP 1:20000; Proteintech), and WT1 (ab15249; 1:800; Abcam). For DNA repair foci experiments, rabbit anti-human RAD51 (ab133534, Abcam) and rabbit anti-human γH2AX antibody (Phospho-Histone H2A.X Ser139; clone 20E3, #9718, Cell Signaling Technologies) were used. For BRCA1 immunoblotting, the mouse monoclonal antibody MS110 antibody was used (Calbiochem OP92).

### Study approvals

For Walter and Eliza Hall Institute of Medical Research (WEHI) PDX models, all experiments involving animals were performed according to the National Health and Medical Research Council Australian Code for the Care and Use of Animals for Scientific Purposes 8th Edition, 2013 (updated 2021), and were approved by the WEHI Animal Ethics Committee (2019.024). Ovarian carcinoma/carcinosarcoma PDX were generated from patients enrolled in the Australian Ovarian Cancer Study (AOCS) or WEHI Stafford Fox Rare Cancer Program (SFRCP). Informed written consent was obtained from all patients, and all experiments were performed according to human ethics guidelines. Additional ethics approval was obtained from the Human Research Ethics Committees at the Royal Women’s Hospital, the WEHI (HREC #10/05 and #G16/02) and QIMR Berghofer (P3456 and P2095). Mouse experiments conducted at FCCC were approved by the Fox Chase Cancer Center Institutional Animal Care and Use Committee (IACUC) and the use of PDX models approved by the Institutional Review Board (IRB).

### Establishment of PDX models

WEHI PDX models were established by transplanting fresh fragments, ascites tumor cells or viably frozen minced tumor (lineage #56C) subcutaneously into NOD/SCID IL2Rγnull recipient mice (T1, passage 1) as described previously (56). Patient details are provided in Supplementary Information. FCCC cohort PDX models #124 and #196 were established as described previously (57) and obtained from Violeta Serra (57). PDX #1126 was established as previously described and obtained from Elgene Lim (41). PDX #204 and FCCC #56 were established by transplanting viable minced tumor into NOD.Cg*-Prkdc^scid^ Il2rg^tm1Wjl^/*SzJ (NSG) recipient mice.

### Treatment of WEHI and FCCC PDX

WEHI cohort recipient mice bearing T2-T10 (passage 2 to passage 10) tumors were randomly assigned to treatments when tumor volume reached 180-300 mm^3^. *In vivo* cisplatin treatments were administered on days 1, 8 and 18, as previously described (34). The regimen for rucaparib treatment was oral gavage once daily Monday-Friday for 3 weeks at 300 mg/kg or 450 mg/kg for all models. Tumors were harvested once tumor volume reached 700 mm^3^ or when mice reached ethical or end of experiment (120 days post treatment) endpoints. Nadir, time to progression (TTP or PD), time to harvest (TTH), and treatment responses are as defined previously (7). Data was plotted using the SurvivalVolume package (58).

Mice bearing PDX tumors from the FCCC cohort were assigned randomly into treatment groups and treatment initiated when tumor volumes reached 150-200 mm^3^. For cisplatin treated mice, a single dose was administered at 6 mg/kg. Rucaparib treatments were administered twice daily Monday-Friday for 2 weeks at 150 mg/kg. Tumors were harvested at 1500 mm^3^ or when the ethical endpoint was reached. Data was plotted using the SurvivalVolume package (58).

### BRCA1 isoform and ABCB1 qRT-PCRs

High quality RNA was extracted from cell pellets or tissue lysates using the Direct-zol RNA MiniPrep (Zymo Research, Cat# R2050) according to the manufacturer’s protocol. Resulting RNA was quantitated using the NanoDrop 2000 UV-Vis Spectrophotometer (Thermo Fisher Scientific, Cat# ND-2000). SuperScript III Reverse Transcriptase (Invitrogen, Cat# 18080085) was used to convert RNA to cDNA according to the manufacturer’s protocol. A volume of 2 ul cDNA diluted to 5 ng/uLng was added to 5 uLul SYBR Green PCR Master Mix (Thermo Fisher Scientific), 2.5 uLul molecular-grade H2O and 0.5 uLul of the relevant 2 uMuM primer mix (Supplementary Information Table 1). *ABCB1* primers used were as previously published (20). Cycling conditions are provided in Supplementary Information.

### Droplet digital PCR assay for WEHI PDX #56PP BRCA1 deletion detection

Droplet digital PCR was performed using the BioRad QX200 Droplet Digital PCR System. PCR reactions were prepared using the QX200 ddPCR EvaGreen Supermix (2X) (BioRad, Catalog #1864034) according to manufacturer’s instructions. PCR details are provided in Supplementary Information.

### Targeted amplicon sequencing of WEHI PDX #049 BRCA1 splice site mutation

PCR reactions were prepared using HotStarTaq DNA Polymerase (QIAGEN, Catalog #203205) according to manufacturer’s instructions, using the following primers for the first step PCR (Illumina Nextera adapters sequences in bold):

Forward primer 5’-3’: **TCGTCGGCAGCGTCAGATGTGTATAAGAGACAGGTGCTCCCCAAAAGCATAA** A;

Reverse primer 5’-3’: **GTCTCGTGGGCTCGGAGATGTGTATAAGAGACAG**GTCTGAAAGCCAGGGAGT TG.

Details of library preparation and sequencing can be found in Supplementary Information.

### SNP arrays and HRD scores

SNP arrays and HRD scores of cell lines and PDX samples were performed as previously described (17).

### DNA sequencing of PDX

BROCA panel sequencing was performed as previously (59) using the BROCA-HR v8 and BROCA-GO v1 versions of the gene panel. For whole exome sequencing, 150-300 ng of DNA was fragmented to approximately 200 bp using a focal acoustic device (Covaris S2, Sage Sciences). Libraries were prepared with the Kapa Hyper Prep Kit (Kapa Biosystems) and SureSelectXT adaptors (Agilent). Hybridization capture was performed with SureSelect Clinical Research Exome V2 baits following the SureSelectXT recommended protocol (Agilent). Indexed libraries were sequenced on an Illumina NextSeq500 to generate on average 130 million paired-end 75 bp reads per sample.

### Whole exome sequencing of cell lines

Library preparation was performed by the Victorian Clinical Genetics Services (VCGS) using the Twist Alliance VCGS Exome custom kit and sequencing was run on the Illumina NovaSeq 6000 using 2×150 bp reads.

### Whole Exome Sequencing Analysis

A bionix (60) pipeline was used to process samples from sequencing data to variant calls. Sequences were aligned to Genome Reference Consortium Human Build 38 (GRCh38) using minimap2 v2.17 (61), and also to *Mus Musculus* reference GRCm38. Mouse derived sequences were removed with XenoMapper v1.0.2 (62). PDX whole exome sequencing used the Agilent SureSelect Clinical Research Exome V2, with reads filtered to 100bp each side of capture regions. Small mutations were called using Octopus v0.7.0 (63) and annotated using SnpEff v4.3 (64). Copy Number Variation was estimated using FACETS v0.6.1 (65).

### RNA sequencing and Analysis

RNA sequencing was performed at the Australian Genomics Research Facility (AGRF) using the Illumina TruSeq Stranded mRNA library preparation, and sequencing was performed using the NovaSeq (SP Lane) at 300 cycles. Reads were mapped using HISAT2 and sashimi plots were visualized in IGV with a threshold of 5 supporting reads.

### Immunohistochemistry protocols

For WEHI PDX models, automated immunohistochemical (IHC) staining was performed on the DAKO Omnis (Agilent Pathology Solutions) samples to confirm ovarian carcinoma/carcinosarcoma characteristics of each PDX, as previously described (17).

### BRCA1 Immunoblotting

Tumor samples were minced and dissociated using a glass homogenizer. Nuclear extracts from the dissociated tumor tissue were obtained using the NE-PER Nuclear and Cytoplasmic Extraction Kit (Thermo Fisher Scientific). Proteins were separated by SDS-PAGE and transferred to a PVDF membrane. Membranes were blocked with 5% nonfat milk in phosphate-buffered saline with Tween 20 (PBST) for 1 hour at room temperature. Primary antibodies were incubated at 4 degrees overnight. Secondary antibodies were incubated for 1 hour at room temperature.

### Cell culture

PEO1, PEO4 and COV362 cell lines were grown at 37°C in 5% CO2 in FV media as previously described (7). UWB1.289 and UWB1.289+BRCA1 cell lines were grown in 50% RPMI 1640 Medium, GlutaMAX™ supplement (Gibco, Cat# 61870127) with 50% MEGM (Mammary Epithelial Growth Medium (MEGM Bullet Kit; Clonetics/Lonza, Cat# CC-3150), and 3% Fetal Bovine Serum (Sigma-Aldrich, Cat #F9423) added. SUM149PT cell lines were grown in Ham’s F-12 with 5 ug/ml insulin, 1 ug/ml hydrocortisone, 10 mM HEPES, 10% FBS, and 1x pen/strep. HEK293T cells were grown in DMEM with 10% FBS. Cell lines confirmed to be negative for Mycoplasma during culture using the MycoAlert Mycoplasma Detection Kit (Lonza, Cat# LT07-118). Whole exome sequencing was used to confirm identity of cell lines.

### BRCA1 minigene experiments

*BRCA1* minigene experiments were performed as previously described (11). Briefly, SSM and primary mutations were introduced into the HA-tagged minigene construct using the Agilent QuickChange Lightning Site-Directed Mutagenesis Kit. Mutant constructs were transfected into HEK293T using the TransIT-LT1 transfection reagent (Mirus). Cell pellets were collected after 72 hours (h) for protein and RNA analysis. Protein expression was assessed by immunoblotting of cell lysates using an anti-HA antibody (Cell Signaling Technology #2367).

### Ectopic BRCA1 expression in PDX

As previously described, BRCA1-Δ11q and mCherry were shuttled into pLENTI-IRES-GFP using Gateway cloning (11). To express ectopic BRCA1-Δ11q and mCherry in PDX #1126, fresh tumor tissue was first extracted and dissociated by shaking for 3 h at 37°C in F-12 media containing 1.6 mg/mL collagenase and 1.6 mg/mL dispase followed by four washes with F-12 media. Dissociated PDX cells were cultured overnight in F-12 media with lentivirus encoding BRCA1-Δ11q or mCherry. Cells were washed, sorted for GFP positivity using a BD-FACS Aria II cell sorter then implanted into NSG mice with an equal ratio of cell suspension and Matrigel (Corning).

### Generation of resistant PDX lineages

Resistant PDX lineages were derived using serial treatments and tumor passaging, summarized schematically in Figure 4H. Tumors were treated with either 6 mg/kg cisplatin or 150 mg/kg rucaparib after reaching 150-200 mm^3^. Rucaparib was administered twice daily Monday-Friday for two weeks. cisplatin was administered at two-week intervals up to four doses if tumors were at least 150 mm^3^. Mice were monitored daily and tumors measured 2-3 times per week. Tumors were collected after reaching 1000 mm^3^ or humane end point for the mice. Fresh collected PDX were reimplanted into NSG mice for additional treatment cycles until tumors stopped responding to treatment or experiment concluded. Cisplatin and rucaparib responses were then assessed as described above for the other PDX models.

### CTG assays

Cells were plated in CELLSTAR® 96 well plates(Greiner, Cat# M1062), at densities related to their growth rates: 500 cells per well for PEO1, 1000 cells for COV362 and 1500 cells per well for PEO4. Drugs were added the following day at the given concentrations (for a total volume of 150 µL per well), and cells were incubated at 37°C in 5% CO2 for 7 days. CellTiter-

Glo® Luminescent (CTG) Cell Viability Assay (Promega, Cat# G7572) was then used to measure viability of cells.

### Cell line gene silencing experiments

Reverse transfection was performed using 1 µL ulLipofectamine™ RNAiMAX Transfection Reagent (Invitrogen, Cat# 13778075) per 100 µLul of Opti-MEM™ I Reduced Serum Medium (Gibco, Cat# 31985062), incubated with 30 nMnM of siRNA for 15-20 minutes before addition of 250,000 cells in culture media (without antibiotics) per well on a 6-well plate. Cells were then either incubated for 2 days before re-seeding for colony formation assays.

Cells plated for colony formation were treated with varying doses of rucaparib the day after re-seeding. Colonies were fixed and stained in 1 x Crystal Violet 0.5% Aqueous Solution (Hurst Scientific, Cat# CV.5-1L) with 20% methanol for 20 minutes. PEO1 cells were fixed at 8 days following treatment, while PEO4 and COV362 cells were fixed at two weeks (accounting for different growth rates). Colonies were counted using the CFU.ai application (http://www.cfu.ai/) and data plotted in PRISM7 (GraphPad).

### Immunofluorescent staining for DNA repair foci

10,000 cells were seeded per well on black PhenoPlate™ 96-well microplates (Perkin Elmer Cat# 6055302) and left to settle for 2 days, before being exposed to 10 Gy of gamma irradiation or left untreated (controls). The Click-iT™ EdU Cell Proliferation Kit for Imaging (Alexa Fluor™ 555 dye) was used to measure cycling cells according to manufacturer’s protocols. Cells were then stained for RAD51 and γH2AX foci and imaged on the Perkin-Elmer OPERA PHENIX platform as previously described (17).

### Sequencing of ARIEL2 and ARIEL4 tumor and plasma DNA samples

DNA, tumor or plasma samples collected from patients in the ARIEL2 (NCT01891344) and ARIEL4 (NCT02855944) clinical trials was sent for gene panel sequencing at Guardant Health, using their FDA-approved TissueNext or Guardant360 CDx tests, respectively.

### Modeling secondary BRCA1 splice mutations

All *in silico* splicing predictions were obtained using NNSPLICE and MaxEnt web tools as previously described (66,67) and SpliceAI with 0.20 threshold as recommended (68). Additional details provided in Supplementary Information.

### BRCA1 isoform competition experiments in SUM149 cells

*BRCA1* exon 11 mutated SUM149 cells were previously engineered using CRISPR/Cas9 to generate a BRCA1-null clone using a single-guide RNA (sgRNA) targeting exon 22 and a full-length expressing BRCA1 clone using an exon 11 targeted sgRNA that introduced a frame-restoring mutation (11). Control BRCA1-Δ11q expressing SUM149 cells were generated using a GFP-targeted sgRNA. One million BRCA1-null cells were premixed with 1,000 cells expressing BRCA1-Δ11q and 1,000 full length BRCA1-expressing cells and injected subcutaneously into the flank NSG mice with an equal volume of Matrigel (Corning). Pelleted cells were also collected for sequencing analysis. Once tumors reached 150-200 mm^3^ mice were subjected to vehicle or cisplatin treatments (6 mg/kg) and tumors collected at 1000 mm^3^. Tumor DNA was extracted and amplicon libraries constructed for the sgRNA sequences using: *forward primer* AATGATACGGCGACCACCGAGATCTACACTCTTTCCCTACACGACGCTCTTCCGATCTTTGTGGAAAGGACGAAACACCG (with additional staggered primers) *barcoded reverse primers* CAAGCAGAAGACGGCATACGAGAT-(8nt index) -GTGACTGGAGTTCAGACGTG TGCTCTTCCGATCTGCCAAGTTGATAA CGGACTAGCCTT. Libraries were sequenced using an Illumina NextSeq2000 and ratios of BRCA1-null, Δ11q, and full length expressing cells determined based on sgRNA content.

### Statistical Analyses

*P* values for PDX model treatments compared to vehicle treatment group were calculated by a log-rank test between fitted Kaplan-Meier estimates with a chi-squared null distribution and one degree of freedom using SurvivalVolumes v1.2.4 (58). *P* values for gene expression differences between PDX models and PDX groups, and for COV362 RAD51 foci analysis, were generated in GraphPad PRISM software using an unpaired t-test with Welch’s correction. *P* < 0.05 was considered statistically significant and statistical tests are indicated in the figure legends. Asterisks indicate statistically significant *P* values.

## Data Availability

All data produced in the present study are available upon reasonable request to the authors.

## Consortia

Australian Ovarian Cancer Study (AOCS), G. Chenevix-Trench, A. Green, P. Webb, D. Gertig, S. Fereday, S. Moore, J. Hung, K. Harrap, T. Sadkowsky, N. Pandeya, M. Malt, A. Mellon, R. Robertson, T. Vanden Bergh, M. Jones, P. Mackenzie, J. Maidens, K. Nattress, Y. E. Chiew, A. Stenlake, H. Sullivan, B. Alexander, P. Ashover, S. Brown, T. Corrish, L. Green, L. Jackman, K. Ferguson, K. Martin, A. Martyn, B. Ranieri, J. White, V. Jayde, P. Mamers, L. Bowes, L. Galletta, D. Giles, J. Hendley, T. Schmidt, H. Shirley, C. Ball, C. Young, S. Viduka, H. Tran, S. Bilic, L. Glavinas, J. Brooks, R. Stuart-Harris, F. Kirsten, J. Rutovitz, P. Clingan, A. Glasgow, A. Proietto, S. Braye, G. Otton, J. Shannon, T. Bonaventura, J. Stewart, S. Begbie, M. Friedlander, D. Bell, S. Baron-Hay, A. Ferrier, G. Gard, D. Nevell, N. Pavlakis, S. Valmadre, B. Young, C. Camaris, R. Crouch, L. Edwards, N. Hacker, D. Marsden, G. Robertson, P. Beale, J. Beith, J. Carter, C. Dalrymple, R. Houghton, P. Russell, M. Links, J. Grygiel, J. Hill, A. Brand, K. Byth, R. Jaworski, P. Harnett, R. Sharma, G. Wain, B. Ward, D. Papadimos, A. Crandon, M. Cummings, K. Horwood, A. Obermair, L. Perrin, D. Wyld, J. Nicklin, M. Davy, M. K. Oehler, C. Hall, T. Dodd, T. Healy, K. Pittman, D. Henderson, J. Miller, J. Pierdes, P. Blomfield, D. Challis, R. McIntosh, A. Parker, B. Brown, R. Rome, D. Allen, P. Grant, S. Hyde, R. Laurie, M. Robbie, D. Healy, T. Jobling, T. Manolitsas, J. McNealage, P. Rogers, B. Susil, E. Sumithran, I. Simpson, K. Phillips, D. Rischin, S. Fox, D. Johnson, S. Lade, M. Loughrey, N. O’Callaghan, W. Murray, P. Waring, V. Billson, J. Pyman, D. Neesham, M. Quinn, C. Underhill, R. Bell, L. F. Ng, R. Blum, V. Ganju, I. Hammond, Y. Leung, A. McCartney, M. Buck, I. Haviv, D. Purdie, D. Whiteman & N. Zeps

## Acknowledgments

We thank Silvia Stoev, Rachel Hancock, Kathy Barber, Scott Wood and Conrad Leonard for technical assistance. We thank Amanda Spurdle and Michael Parsons (QIMR Berghofer) for their advice regarding *BRCA1* SNPs potentially affecting splicing. We thank Clovis Oncology for providing rucaparib for *in vivo* experiments; Violeta Serra and Judith Balmana for the PDX models #124 and #196; Elgene Lim for #1126, Scott H. Kaufmann for PEO1 and PEO4 cell lines and Dale Garsed and David Bowtell for the UWB1.289 cell line. This work was supported by fellowships and grants from the National Health and Medical Research Council (NHMRC Australia; Project grant 1062702 (CLS); Investigator grant 2009783 (CLS)); the Stafford Fox Medical Research Foundation (CLS); Cancer Council Victoria (Sir Edward Dunlop Fellowship in Cancer Research (CLS)); the Victorian Cancer Agency (Clinical Fellowships CRF10–20, CRF16014 (CSL)); the National Institute of Health (2P50CA083636) (to EMS), NIH NCI R01 CA251817 and NIH NCI R01 CA237600 (to EMS and MR), and the Wendy Feuer Ovarian Cancer Research Fund (to EMS); the Bev Gray Ovarian Cancer Scholarship (PhD top-up scholarship) and Research Training Program Scholarship (PhD Scholarship) (KN); the American Association of Cancer Research (AACR-AstraZeneca Ovarian Cancer Research Fellowship 2022 (KN)); the Swiss Cancer Research foundation (KFS-5445-08-2021) (FG); NHMRC Emerging Leader 1 Investigator Grant (APP2008631) (OK), and Stand Up to Cancer (EMS). This work was supported by US National Institutes of Health (NIH) Grants R01CA214799 and R01GM135293 to NJ. J.J.K. was supported by an American Cancer Society—Tri State CEOs Against Cancer Postdoctoral Fellowship, PF-19-097-01-DMC, Ovarian Cancer Research Alliance and Phil and Judy Messing grant 597484. This work was made possible through the Australian Cancer Research Foundation, the Victorian State Government Operational Infrastructure Support and Australian Government NHMRC IRIISS. All authors, the WEHI Stafford Fox Rare Cancer Program and the AOCS would like to thank all of the patients who participated in these research programs. The AOCS would also like to acknowledge the contribution of the study nurses, research assistants, and all clinical and scientific collaborators to the study. The complete AOCS Study Group can be found at www.aocstudy.org. The Australian Ovarian Cancer Study gratefully acknowledges additional support from Ovarian Cancer Australia and the Peter MacCallum Foundation. The Australian Ovarian Cancer Study Group was supported by the U.S. Army Medical Research and Materiel Command under DAMD17-01-1-0729, The Cancer Council Victoria, Queensland Cancer Fund, The Cancer Council New South Wales, The Cancer Council South Australia, The Cancer Council Tasmania and The Cancer Foundation of Western Australia (Multi-State Applications 191, 211 and 182) and the National Health and Medical Research Council of Australia (NHMRC; ID199600; ID400413 and ID400281)

## References

1. Moore K, Colombo N, Scambia G, Kim BG, Oaknin A, Friedlander M, et al. Maintenance Olaparib in Patients with Newly Diagnosed Advanced Ovarian Cancer. N Engl J Med 2018;379(26):2495–505 doi 10.1056/NEJMoa1810858.

2. González-Martín A, Pothuri B, Vergote I, DePont Christensen R, Graybill W, Mirza MR, et al. Niraparib in Patients with Newly Diagnosed Advanced Ovarian Cancer. New England Journal of Medicine 2019;381(25):2391–402 doi 10.1056/NEJMoa1910962.

3. Ray-Coquard I, Pautier P, Pignata S, Pérol D, González-Martín A, Berger R, et al. Olaparib plus Bevacizumab as First-Line Maintenance in Ovarian Cancer. New England Journal of Medicine 2019;381(25):2416–28.

4. Sakai W, Swisher EM, Karlan BY, Agarwal MK, Higgins J, Friedman C, et al. Secondary mutations as a mechanism of cisplatin resistance in BRCA2-mutated cancers. Nature 2008;451(7182):1116–20 doi 10.1038/nature06633.

5. Edwards SL, Brough R, Lord CJ, Natrajan R, Vatcheva R, Levine DA, et al. Resistance to therapy caused by intragenic deletion in BRCA2. Nature 2008;451(7182):1111–5 doi 10.1038/nature06548.

6. Patch AM, Christie EL, Etemadmoghadam D, Garsed DW, George J, Fereday S, et al. Whole-genome characterization of chemoresistant ovarian cancer. Nature 2015;521(7553):489-94 doi 10.1038/nature14410.

7. Kondrashova O, Topp M, Nesic K, Lieschke E, Ho GY, Harrell MI, et al. Methylation of all BRCA1 copies predicts response to the PARP inhibitor rucaparib in ovarian carcinoma. Nat Commun 2018;9(1):3970 doi 10.1038/s41467-018-05564-z.

8. Sakai W, Swisher EM, Jacquemont C, Chandramohan KV, Couch FJ, Langdon SP, et al. Functional restoration of BRCA2 protein by secondary BRCA2 mutations in BRCA2-mutated ovarian carcinoma. Cancer Res 2009;69(16):6381–6 doi 10.1158/0008-5472.CAN-09-1178.

9. Swisher EM, Sakai W, Karlan BY, Wurz K, Urban N, Taniguchi T. Secondary BRCA1 mutations in BRCA1-mutated ovarian carcinomas with platinum resistance. Cancer Res 2008;68(8):2581–6 doi 10.1158/0008-5472.Can-08-0088.

10. Johnson N, Johnson SF, Yao W, Li YC, Choi YE, Bernhardy AJ, et al. Stabilization of mutant BRCA1 protein confers PARP inhibitor and platinum resistance. Proc Natl Acad Sci U S A 2013;110(42):17041–6 doi 10.1073/pnas.1305170110.

11. Wang Y, Bernhardy AJ, Cruz C, Krais JJ, Nacson J, Nicolas E, et al. The BRCA1-Delta11q Alternative Splice Isoform Bypasses Germline Mutations and Promotes Therapeutic Resistance to PARP Inhibition and Cisplatin. Cancer Res 2016;76(9):2778–90 doi 10.1158/0008-5472.CAN-16-0186.

12. Bouwman P, Aly A, Escandell JM, Pieterse M, Bartkova J, van der Gulden H, et al. 53BP1 loss rescues BRCA1 deficiency and is associated with triple-negative and BRCA-mutated breast cancers. Nat Struct Mol Biol 2010;17(6):688–95 doi 10.1038/nsmb.1831.

13. Bunting SF, Callen E, Wong N, Chen HT, Polato F, Gunn A, et al. 53BP1 inhibits homologous recombination in Brca1-deficient cells by blocking resection of DNA breaks. Cell 2010;141(2):243–54 doi 10.1016/j.cell.2010.03.012.

14. Gupta R, Somyajit K, Narita T, Maskey E, Stanlie A, Kremer M, et al. DNA Repair Network Analysis Reveals Shieldin as a Key Regulator of NHEJ and PARP Inhibitor Sensitivity. Cell 2018;173(4):972–88.e23 doi 10.1016/j.cell.2018.03.050.

15. Dev H, Chiang T-WW, Lescale C, de Krijger I, Martin AG, Pilger D, et al. Shieldin complex promotes DNA end-joining and counters homologous recombination in BRCA1-null cells. Nature cell biology 2018;20(8):954–65.

16. Pettitt SJ, Krastev DB, Brandsma I, Drean A, Song F, Aleksandrov R, et al. Genome-wide and high-density CRISPR-Cas9 screens identify point mutations in PARP1 causing PARP inhibitor resistance. Nat Commun 2018;9(1):1849 doi 10.1038/s41467-018-03917-2.

17. Nesic K, Kondrashova O, Hurley RM, McGehee CD, Vandenberg CJ, Ho G-Y, et al. Acquired RAD51C promoter methylation loss causes PARP inhibitor resistance in high-grade serous ovarian carcinoma. Cancer research 2021;81(18):4709–22.

18. Swisher EM, Kwan TT, Oza AM, Tinker AV, Ray-Coquard I, Oaknin A, et al. Molecular and clinical determinants of response and resistance to rucaparib for recurrent ovarian cancer treatment in ARIEL2 (Parts 1 and 2). Nat Commun 2021;12(1):2487 doi 10.1038/s41467-021-22582-6.

19. Cong K, Peng M, Kousholt AN, Lee WTC, Lee S, Nayak S, et al. Replication gaps are a key determinant of PARP inhibitor synthetic lethality with BRCA deficiency. Mol Cell 2021;81(15):3128–44.e7 doi 10.1016/j.molcel.2021.06.011.

20. Christie EL, Pattnaik S, Beach J, Copeland A, Rashoo N, Fereday S, et al. Multiple ABCB1 transcriptional fusions in drug resistant high-grade serous ovarian and breast cancer. Nature Communications 2019;10(1):1295 doi 10.1038/s41467-019-09312-9.

21. Dimitrova D, Ruscito I, Olek S, Richter R, Hellwag A, Türbachova I, et al. Germline mutations of BRCA1 gene exon 11 are not associated with platinum response neither with survival advantage in patients with primary ovarian cancer: understanding the clinical importance of one of the biggest human exons. A study of the Tumor Bank Ovarian Cancer (TOC) Consortium. Tumour Biol 2016;37(9):12329–37 doi 10.1007/s13277-016-5109-8.

22. Orban TI, Olah E. Expression profiles of BRCA1 splice variants in asynchronous and in G1/S synchronized tumor cell lines. Biochem Biophys Res Commun 2001;280(1):32–8 doi 10.1006/bbrc.2000.4068.

23. Raponi M, Douglas AG, Tammaro C, Wilson DI, Baralle D. Evolutionary constraint helps unmask a splicing regulatory region in BRCA1 exon 11. PLoS One 2012;7(5):e37255 doi 10.1371/journal.pone.0037255.

24. Tammaro C, Raponi M, Wilson DI, Baralle D. BRCA1 exon 11 alternative splicing, multiple functions and the association with cancer. Biochem Soc Trans 2012;40(4):768–72 doi 10.1042/bst20120140.

25. Ludwig T, Chapman DL, Papaioannou VE, Efstratiadis A. Targeted mutations of breast cancer susceptibility gene homologs in mice: lethal phenotypes of Brca1, Brca2, Brca1/Brca2, Brca1/p53, and Brca2/p53 nullizygous embryos. Genes Dev 1997;11(10):1226–41 doi 10.1101/gad.11.10.1226.

26. Xu X, Wagner KU, Larson D, Weaver Z, Li C, Ried T, et al. Conditional mutation of Brca1 in mammary epithelial cells results in blunted ductal morphogenesis and tumour formation. Nat Genet 1999;22(1):37–43 doi 10.1038/8743.

27. Xu X, Qiao W, Linke SP, Cao L, Li WM, Furth PA, et al. Genetic interactions between tumor suppressors Brca1 and p53 in apoptosis, cell cycle and tumorigenesis. Nat Genet 2001;28(3):266–71 doi 10.1038/90108.

28. Cao L, Li W, Kim S, Brodie SG, Deng CX. Senescence, aging, and malignant transformation mediated by p53 in mice lacking the Brca1 full-length isoform. Genes Dev 2003;17(2):201–13 doi 10.1101/gad.1050003.

29. Perrin-Vidoz L, Sinilnikova OM, Stoppa-Lyonnet D, Lenoir GM, Mazoyer S. The nonsense-mediated mRNA decay pathway triggers degradation of most BRCA1 mRNAs bearing premature termination codons. Hum Mol Genet 2002;11(23):2805–14 doi 10.1093/hmg/11.23.2805.

30. Breast Cancer Information Core. <https://research.nhgri.nih.gov/projects/bic/index.shtml>.

31. Thompson D, Easton D. Variation in BRCA1 cancer risks by mutation position. Cancer Epidemiol Biomarkers Prev 2002;11(4):329–36.

32. Risch HA, McLaughlin JR, Cole DE, Rosen B, Bradley L, Kwan E, et al. Prevalence and penetrance of germline BRCA1 and BRCA2 mutations in a population series of 649 women with ovarian cancer. Am J Hum Genet 2001;68(3):700–10 doi 10.1086/318787.

33. Risch HA, McLaughlin JR, Cole DE, Rosen B, Bradley L, Fan I, et al. Population BRCA1 and BRCA2 mutation frequencies and cancer penetrances: a kin-cohort study in Ontario, Canada. J Natl Cancer Inst 2006;98(23):1694–706 doi 10.1093/jnci/djj465.

34. Topp MD, Hartley L, Cook M, Heong V, Boehm E, McShane L, et al. Molecular correlates of platinum response in human high-grade serous ovarian cancer patient-derived xenografts. Mol Oncol 2014;8(3):656–68 doi 10.1016/j.molonc.2014.01.008.

35. Wappenschmidt B, Becker AA, Hauke J, Weber U, Engert S, Köhler J, et al. Analysis of 30 putative BRCA1 splicing mutations in hereditary breast and ovarian cancer families identifies exonic splice site mutations that escape in silico prediction. PLoS One 2012;7(12):e50800 doi 10.1371/journal.pone.0050800.

36. Bonatti F, Pepe C, Tancredi M, Lombardi G, Aretini P, Sensi E, et al. RNA-based analysis of BRCA1 and BRCA2 gene alterations. Cancer Genet Cytogenet 2006;170(2):93–101 doi 10.1016/j.cancergencyto.2006.05.005.

37. Hurley RM, Wahner Hendrickson AE, Visscher DW, Ansell P, Harrell MI, Wagner JM, et al. 53BP1 as a potential predictor of response in PARP inhibitor-treated homologous recombination-deficient ovarian cancer. Gynecol Oncol 2019;153(1):127–34 doi 10.1016/j.ygyno.2019.01.015.

38. Haley J, Tomar S, Pulliam N, Xiong S, Perkins SM, Karpf AR, et al. Functional characterization of a panel of high-grade serous ovarian cancer cell lines as representative experimental models of the disease. Oncotarget 2016;7(22):32810–20 doi 10.18632/oncotarget.9053.

39. Ruiz de Garibay G, Fernandez-Garcia I, Mazoyer S, Leme de Calais F, Ameri P, Vijayakumar S, et al. Altered regulation of BRCA1 exon 11 splicing is associated with breast cancer risk in carriers of BRCA1 pathogenic variants. Hum Mutat 2021;42(11):1488–502 doi 10.1002/humu.24276.

40. Anczuków O, Buisson M, Salles MJ, Triboulet S, Longy M, Lidereau R, et al. Unclassified variants identified in BRCA1 exon 11: Consequences on splicing. Genes Chromosomes Cancer 2008;47(5):418–26 doi 10.1002/gcc.20546.

41. Johnson SF, Cruz C, Greifenberg AK, Dust S, Stover DG, Chi D, et al. CDK12 Inhibition Reverses De Novo and Acquired PARP Inhibitor Resistance in BRCA Wild-Type and Mutated Models of Triple-Negative Breast Cancer. Cell Reports 2016;17(9):2367–81 doi 10.1016/j.celrep.2016.10.077.

42. Arason A, Agnarsson BA, Johannesdottir G, Johannsson OT, Hilmarsdottir B, Reynisdottir I, et al. The BRCA1 c.4096+3A>G Variant Displays Classical Characteristics of Pathogenic BRCA1 Mutations in Hereditary Breast and Ovarian Cancers, But Still Allows Homozygous Viability. Genes (Basel) 2019;10(11):882 doi 10.3390/genes10110882.

43. Slavin TP, Manjarrez S, Pritchard CC, Gray S, Weitzel JN. The effects of genomic germline variant reclassification on clinical cancer care. Oncotarget 2019;10(4).

44. Byrjalsen A, Steffensen AY, Hansen TVO, Wadt K, Gerdes A-M. Classification of the spliceogenic BRCA1 c.4096+3A>G variant as likely benign based on cosegregation data and identification of a healthy homozygous carrier. Clin Case Rep 2017;5(6):876–9 doi 10.1002/ccr3.944.

45. Kondrashova O, Nguyen M, Shield-Artin K, Tinker AV, Teng NNH, Harrell MI, et al. Secondary Somatic Mutations Restoring RAD51C and RAD51D Associated with Acquired Resistance to the PARP Inhibitor Rucaparib in High-Grade Ovarian Carcinoma. Cancer Discov 2017;7(9):984–98 doi 10.1158/2159-8290.CD-17-0419.

46. van den Berg-Bakker CA, Hagemeijer A, Franken-Postma EM, Smit VT, Kuppen PJ, van Ravenswaay Claasen HH, et al. Establishment and characterization of 7 ovarian carcinoma cell lines and one granulosa tumor cell line: growth features and cytogenetics. Int J Cancer 1993;53(4):613–20 doi 10.1002/ijc.2910530415.

47. Domcke S, Sinha R, Levine DA, Sander C, Schultz N. Evaluating cell lines as tumour models by comparison of genomic profiles. Nat Commun 2013;4:2126 doi 10.1038/ncomms3126.

48. Barnes BM, Nelson L, Tighe A, Morgan RD, McGrail J, Taylor SS. Classification of ovarian cancer cell lines using transcriptional profiles defines the five major pathological subtypes. bioRxiv 2020:2020.07.14.202457 doi 10.1101/2020.07.14.202457.

49. Fraile-Bethencourt E, Valenzuela-Palomo A, Díez-Gómez B, Goina E, Acedo A, Buratti E, et al. Mis-splicing in breast cancer: identification of pathogenic BRCA2 variants by systematic minigene assays. J Pathol 2019;248(4):409–20 doi 10.1002/path.5268.

50. Nacson J, Krais JJ, Bernhardy AJ, Clausen E, Feng W, Wang Y, et al. BRCA1 Mutation-Specific Responses to 53BP1 Loss-Induced Homologous Recombination and PARP Inhibitor Resistance. Cell Reports 2018;24(13):3513–27.e7 doi 10.1016/j.celrep.2018.08.086.

51. Burdett NL, Willis MO, Alsop K, Hunt AL, Pandey A, Hamilton PT, et al. Multiomic analysis of homologous recombination-deficient end-stage high-grade serous ovarian cancer. Nat Genet 2023 doi 10.1038/s41588-023-01320-2.

52. Eskens FALM, Ramos FJ, Burger H, O’Brien JP, Piera A, de Jonge MJA, et al. Phase I Pharmacokinetic and Pharmacodynamic Study of the First-in-Class Spliceosome Inhibitor E7107 in Patients with Advanced Solid Tumors. Clinical Cancer Research 2013;19(22):6296–304 doi 10.1158/1078-0432.Ccr-13-0485.

53. Zhang Y, Qian J, Gu C, Yang Y. Alternative splicing and cancer: a systematic review. Signal Transduction and Targeted Therapy 2021;6(1):78 doi 10.1038/s41392-021-00486-7.

54. Steensma DP, Wermke M, Klimek VM, Greenberg PL, Font P, Komrokji RS, et al. Phase I First-in-Human Dose Escalation Study of the oral SF3B1 modulator H3B-8800 in myeloid neoplasms. Leukemia 2021;35(12):3542–50 doi 10.1038/s41375-021-01328-9.

55. Esposito D, Weile J, Shendure J, Starita LM, Papenfuss AT, Roth FP, et al. MaveDB: an open-source platform to distribute and interpret data from multiplexed assays of variant effect. Genome Biology 2019;20(1):223 doi 10.1186/s13059-019-1845-6.

56. Kuchenbaecker KB, Hopper JL, Barnes DR, Phillips KA, Mooij TM, Roos-Blom MJ, et al. Risks of Breast, Ovarian, and Contralateral Breast Cancer for BRCA1 and BRCA2 Mutation Carriers. JAMA 2017;317(23):2402–16 doi 10.1001/jama.2017.7112.

57. Cruz C, Castroviejo-Bermejo M, Gutiérrez-Enríquez S, Llop-Guevara A, Ibrahim YH, Gris-Oliver A, et al. RAD51 foci as a functional biomarker of homologous recombination repair and PARP inhibitor resistance in germline BRCA-mutated breast cancer. Annals of Oncology 2018;29(5):1203–10 doi 10.1093/annonc/mdy099.

58. Wakefield MJ. SurvivalVolume: interactive volume threshold survival graphs. J Open Source Software 2016;1(8):111.

59. Walsh T, Casadei S, Lee MK, Pennil CC, Nord AS, Thornton AM, et al. Mutations in 12 genes for inherited ovarian, fallopian tube, and peritoneal carcinoma identified by massively parallel sequencing. Proc Natl Acad Sci U S A 2011;108(44):18032–7 doi 10.1073/pnas.1115052108.

60. Bedő J, Di Stefano L, Papenfuss AT. Unifying package managers, workflow engines, and containers: Computational reproducibility with BioNix. GigaScience 2020;9(11) doi 10.1093/gigascience/giaa121.

61. Li H. Minimap2: pairwise alignment for nucleotide sequences. Bioinformatics 2018;34(18):3094–100 doi 10.1093/bioinformatics/bty191.

62. Wakefield MJ. Xenomapper: Mapping reads in a mixed species context. Journal of Open Source Software 2016;1(1):18 doi 10.21105/joss.00018.

63. Cooke DP, Wedge DC, Lunter G. A unified haplotype-based method for accurate and comprehensive variant calling. Nature biotechnology 2021 doi 10.1038/s41587-021-00861-3.

64. Cingolani PaP, A. and Coon, M. and Nguyen, T. and Wang, L. and Land, S.J. and Lu, X. and Ruden, D.M. A program for annotating and predicting the effects of single nucleotide polymorphisms, SnpEff: SNPs in the genome of Drosophila melanogaster strain w1118; iso-2; iso-3. Fly 2012;6(2):13.

65. Shen R, Seshan VE. FACETS: allele-specific copy number and clonal heterogeneity analysis tool for high-throughput DNA sequencing. Nucleic Acids Res 2016;44(16):e131 doi 10.1093/nar/gkw520.

66. Casadei S, Gulsuner S, Shirts BH, Mandell JB, Kortbawi HM, Norquist BS, et al. Characterization of splice-altering mutations in inherited predisposition to cancer. Proc Natl Acad Sci U S A 2019;116(52):26798–807 doi 10.1073/pnas.1915608116.

67. Abu Rayyan A, Kamal L, Casadei S, Brownstein Z, Zahdeh F, Shahin H, et al. Genomic analysis of inherited hearing loss in the Palestinian population. Proc Natl Acad Sci U S A 2020;117(33):20070–6 doi 10.1073/pnas.2009628117.

68. Jaganathan K, Kyriazopoulou Panagiotopoulou S, McRae JF, Darbandi SF, Knowles D, Li YI, et al. Predicting Splicing from Primary Sequence with Deep Learning. Cell 2019;176(3):535–48.e24 doi 10.1016/j.cell.2018.12.015.

