## Supplementary Information for "*BRCA1* secondary splice-site mutations drive exon-skipping and PARP inhibitor resistance"

Nesic et al., 2023

***Detailed WEHI patient clinical information and description of corresponding PDX***

OCS PDX #264 was generated from a stage IIIC ovarian carcinosarcoma (OCS) tumor sample from an individual who was found to be a germline *BRCA1* mutation carrier (also known to have had two TNBC and a melanoma), who had received five lines of prior therapy, including one line of PARPi Lynparza/Olaparib (Table 1). Their HGSOC was initially responsive to platinum chemotherapy, with an initial platinum treatment free interval (TFI) of 15 months and in the relapsed maintenance setting they remained on PARPi for > two years. OCS PDX #264 demonstrated progressive disease on both 4 mg/kg cisplatin, (Time to progression of four days (TTP) for cisplatin and vehicle; p= 0.004; Figure 1A; Supplementary table S2) and on the 450 mg/kg dose of PARPi rucaparib (TTP of 4 days for both rucaparib and vehicle arms; p= 0.502), reflecting the fact that her OCS had progressed and become refractory to both platinum and PARPi in the clinic. OCS PDX #264 harboured a germline exon 11 *BRCA1*: c.2216_2217delAA mutation and a pathogenic *TP53*: p.Glu336* mutation (Table 1) in tumor tissue obtained following five lines of therapy, including two years of PARPi therapy.

HGSOC PDX #56 was derived from an individual diagnosed with stage III HGSOC, who had not had any prior therapy, and the original lineage of this PDX has been published previously (1). HGSOC PDX #56 harboured a germline *BRCA1*: c.894_895delTG mutation, as well as a *TP53*: c.963del mutation and *RB1* truncation. A new lineage of this PDX, derived from a post-cisplatin PDX tumor, is presented in this study: HGSOC PDX #56C (Supplementary Figure S1; Supplementary table S2). Like the original #56 PDX lineage, #56C was found to be cisplatin-sensitive (TTP of 53 days for cisplatin vs four days for vehicle; p= <0.001) and showed some response to the PARPi rucaparib at 300 mg/kg (TTP of 71 days for rucaparib vs four days for vehicle; p= <0.001).

HGSOC PDX #56PP was derived from an inguinal lymph node biopsy from patient #56 following a total of five prior lines of therapy, including 14 months of second-line PARPi in the second line (Table 1). This individual’s HGSOC was initially sensitive to first line carboplatin/paclitaxel, but they developed progressive disease on subsequent lines of therapy (including progressive disease on re-challenge with 7^th^ line PARPi). HGSOC PDX #56PP demonstrated some response to cisplatin, with a delay in median TTP (36 days for cisplatin versus four days for vehicle; p= 0.051; Figure 1A; Supplementary table S2), however it showed no response to the PARPi rucaparib at the high 450 mg/kg dose administered five days a week for three weeks, which reflected the refractory nature of the patient’s HGSOC response to re-challenge with 7^th^ line PARPi (TTP of four days for both vehicle and rucaparib; p= 0.375). HGSOC PDX #56PP carried a germline *BRCA1*: c.895_896del mutation, *TP53*:c.963delA and *PTEN*:c.166delT (Table 1).

HGSOC PDX #032 was generated from an individual diagnosed with recurrent metastatic HGSOC, who was found to have a somatic mutation in *BRCA1* in their cancer, who had received six prior lines of therapy, including PARPi in the 4^th^ line setting (combined with cyclophosphamide in the SOLACE2 clinical trial). This patient had partial sensitivity to 1^st^ line carboplatin/paclitaxel (platinum TFI of 11 months), but relapsed three times prior to receiving PARPi, to which their HGSOC had an initial response followed by progressive disease. HGSOC PDX #032 demonstrated some response to 4mg/kg cisplatin *in vivo* (TTP of 67 days for cisplatin vs eight days for vehicle; p= 0.093; Figure 1A; Supplementary table S2) and was not responsive to rucaparib 450 mg/kg (Supplementary table S2). HGSOC PDX #032, obtained after six lines of therapy including a PARPi, harboured a somatic *BRCA1*: c.1251delT mutation in exon 11, along with deletions in TP53 (c.97-1_97-6del) and FANCA (c.3788_3790del; Table 1).

HGSOC PDX #049 was derived from an individual with HGSOC who was a germline *BRCA1* mutation carrier with a prior TNBC, who had received four lines of chemotherapy prior to PDX establishment, including PARPi maintenance therapy in the 2^nd^ line for 13 months followed by progressive disease on 3rd line cisplatin chemotherapy. HGSOC PDX #049 developed stable disease in response to 4 mg/kg cisplatin treatment (TTP of >120 days for cisplatin arm, compared to four days for vehicle arm; p= 0.01), but was less responsive to 300 mg/kg of PARPi rucaparib (TTP of 22 days for rucaparib vs four days for vehicle arm; p= 0.01). HGSOC PDX #1049, obtained after four lines of therapy, including PARPi, was found to harbour two *BRCA1* mutations – a germline c.2475delC mutation (100% allele frequency) and a TP53 c.428_429insCA mutation.

HGSOC PDX #206 was derived from an individual who was known to belong to a *BRCA1* mutation positive family, who had been diagnosed with stage IV HGSOC and was initially platinum sensitive (TFI of 17 months). They had received one line of carboplatin/paclitaxel chemotherapy prior to PDX establishment. HGSOC PDX #206 was found to be highly sensitive to both cisplatin (TTP of 7 days for vehicle arm vs >120 days for cisplatin arm; p= 0.001) and rucaparib at a 300mg/kg dose (TTP of 7 days for vehicle arm vs >120 days, p= 0.002). HGSOC PDX #206, obtained after one line of carboplatin/paclitaxel, had a germline *BRCA1*: c.3817C>T mutation in exon 11, and a *TP53*:c.451C>T mutation and *BLM* rearrangement. The patient was found to have later developed a reversion mutation in *BRCA1* (to wild-type) from the ALLOCATE study panel sequencing of a post-progression biopsy (post-second-line platinum and olaparib) (2).

***Cycling conditions for BRCA1 isoform and ABCB1 qPCRs***

Plates were incubated on the ABI 7900 (Applied Biosystems) or the ViiA 7 Real-Time PCR System (Thermo Fisher Scientific) at 95°C for 10 minutes, 40 cycles of 95°C for 15 seconds and 60°C for 1 minute*, followed by 95°C for 15 seconds, 60°C for 15 seconds and 95°C for 15 seconds* (*Data recorded at indicated steps). Resulting CT values were normalized to the averaged value housekeeper control primers in Excel (Microsoft). Fold change values were calculated for each sample. These values were then graphed in PRISM7 (GraphPad), and the standard deviation calculated.

***Supplementary Methods Table 1.*** *HPRT, B-ACTIN, SDHA and GAPDH were used as housekeeper control primers. A BRCA1 primer set for all transcripts was used for normalization of each sample to the total amount of all BRCA1 transcripts. BRCA1 +exon 11 primer set specifically amplifies BRCA1 transcripts with all of exon 11. The BRCA1 D11q and D11 primer sets were designed to be specific to their respective isoforms.*

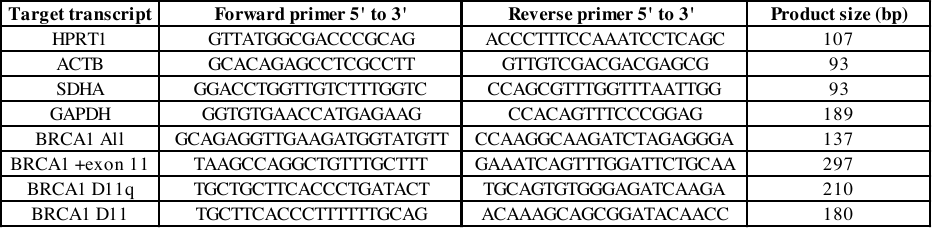

***WES and BROCA analysis for resistance mutations***

In WES data for PDX #032, #049 or #56PP, no variants were detected (adjusted variant allele frequency of >5%) in the following potential *BRCA1*-mutant PARPi-resistance genes: *PARP1, SHLD1 (C20orf196), SHLD2 (FAM35A), SHLD3 (CTC-534A2.2), TP53BP1, RIF1, MAD2L2, REV7, DYNLL1, PTIP* (3-6). PDX #196 and #264 only had BROCA available. PDX #264 was run on the BROCA-GO v1 panel where no mutations in *MAD2L2, PARP1, RIF1, SHLD1, SHLD2, SHLD3* or *TP53BP1* were found. #196 was run on the older BROCA v8 panel, where no mutations were found in *MAD2L2, PARP1, RIF1* or *TP53BP1.*

***Targeted amplicon sequencing of WEHI PDX #049 splice site and TP53 mutation***

PCR reactions were prepared using HotStarTaq DNA Polymerase (QIAGEN, Catalog #203205) according to manufacturer’s instructions. #049 SSM primers are outlined in main methods. TP53 mutation primers for first step PCR (Illumina Nextera adapters sequences in bold):

**TCGTCGGCAGCGTCAGATGTGTATAAGAGACAG**AGCTCGCTAGTGGGTTGCAGGA; Reverse primer 5’-3’:

**GTCTCGTGGGCTCGGAGATGTGTATAAGAGACAG**TGCTTGTAGATGGCCATGGCGC.

Reactions were incubated at 95ºC for 15 minutes, followed by 15 cycles of 95ºC for 30 seconds, 55ºC (SSM reaction) or 65ºC (TP53 reaction) for 40 seconds and 72 ºC for 40 seconds. Reactions were then incubated at 72ºC for 10 minutes and storage at 4ºC. Illumina Nexera XT indexes were then incorporated in a second PCR, as previously described (7). Resulting libraries were cleaned-up using Agencourt AMPure XP (Beckman Coulter, Cat#10136224) beads using a ratio of 0.9:1. The D1000 ScreenTape System (Agilent, Cat#5067-5582 and 5067-5583) was used to assess the size profile of NexteraXT libraries according to manufacturer’s protocol. Libraries were sequenced in the WEHI Genomics Core Laboratory on the Illumina MiSeq using a MiSeq Nano Reagent Kit v2 (300 cycle; Illumina; Cat# MS-102-2002) according to manufacturer’s protocols.

***Droplet digital PCR assay for WEHI PDX #56PP deletion detection***

Two reactions were prepared; one for wildtype allele detection and one for deletion allele detection. Forward primer 5’ TCCCCATCATGTGAGTCATC 3’, and reverse primer 5’ CTGATTGTTGCCCTTTCGTT 3’ were used for deletion allele detection. The same forward primer was used for wildtype alleles, combined with reverse primer 5’ TCTGAATGCTGATCCCCTGT 3’. Reactions were incubated at 95ºC for 5 minutes, followed by 40 cyles of 95ºC for 30 seconds and 60ºC	for 60 seconds. Reactions were then incubated at 4ºC for 5 minutes, 90ºC for 5 minutes and storage at 4ºC.

***In Silico modelling secondary BRCA1 splice mutations***

All *in silico* splicing predictions were obtained using NNSPLICE and MaxEnt web tools as previously described (8,9) and SpliceAI with 0.20 threshold as recommended (10).

For secondary mutations at or near *BRCA1* c.4096 (exon 10), transcript outcomes were modeled upon *BRCA1* c.4096+3A>G at chr17:41,243,449T>C (CF5004.01), which has been shown experimentally to lead to (a) skipping of exon 10 (del 3426bp; del aa 224-1365) in 24% and (b) cryptic splice D11q (del 3309bp; del aa 264-1366) in 70% transcript from the mutant allele (King/Swisher Lab, unpublished data; (11)).

For secondary mutations at or near c.5277 (exon 19), transcript outcomes were modeled upon *BRCA1* c.5277+1G>A at chr17:41,209,068C>T (CF1012.03), which was shown experimentally to lead to (a) skipping of exon 20 (del 84bp; del aa 1732-1759) in 14% and (b) cryptic donor splice in intron 20 (ins 87bp, 1676 stop) in 48% transcript from the mutant allele (8).

For *BRCA1* c.4986+2T>G (exon 16), transcript outcomes were modeled upon *BRCA1* c.4986+3G>C at chr17:41,222,942C>G (CF4616.01), which was shown experimentally to disrupt normal splicing and to lead to a cryptic donor splice in intron 16 (ins 65bp, stop 1676) in 44% transcript from the mutant allele (8,11).

**References**

1.	Kondrashova O, Topp M, Nesic K, Lieschke E, Ho GY, Harrell MI*, et al.* Methylation of all BRCA1 copies predicts response to the PARP inhibitor rucaparib in ovarian carcinoma. Nat Commun **2018**;9(1):3970 doi 10.1038/s41467-018-05564-z.

2.	Kondrashova O, Ho GY, Au-Yeung G, Leas L, Boughtwood T, Alsop K*, et al.* Clinical Utility of Real-Time Targeted Molecular Profiling in the Clinical Management of Ovarian Cancer: The ALLOCATE Study. JCO Precis Oncol **2019**;3:1-18 doi 10.1200/po.19.00019.

3.	He YJ, Meghani K, Caron M-C, Yang C, Ronato DA, Bian J*, et al.* DYNLL1 binds to MRE11 to limit DNA end resection in BRCA1-deficient cells. Nature **2018**;563(7732):522-6 doi 10.1038/s41586-018-0670-5.

4.	Bunting SF, Callen E, Wong N, Chen HT, Polato F, Gunn A*, et al.* 53BP1 inhibits homologous recombination in Brca1-deficient cells by blocking resection of DNA breaks. Cell **2010**;141(2):243-54 doi 10.1016/j.cell.2010.03.012.

5.	Xu G, Chapman JR, Brandsma I, Yuan J, Mistrik M, Bouwman P*, et al.* REV7 counteracts DNA double-strand break resection and affects PARP inhibition. Nature **2015**;521(7553):541-4 doi 10.1038/nature14328.

6.	Ray Chaudhuri A, Callen E, Ding X, Gogola E, Duarte AA, Lee J-E*, et al.* Replication fork stability confers chemoresistance in BRCA-deficient cells. Nature **2016**;535(7612):382-7 doi 10.1038/nature18325.

7.	Nesic K, Kondrashova O, Hurley RM, McGehee CD, Vandenberg CJ, Ho G-Y*, et al.* Acquired RAD51C promoter methylation loss causes PARP inhibitor resistance in high-grade serous ovarian carcinoma. Cancer research **2021**;81(18):4709-22.

8.	Casadei S, Gulsuner S, Shirts BH, Mandell JB, Kortbawi HM, Norquist BS*, et al.* Characterization of splice-altering mutations in inherited predisposition to cancer. Proc Natl Acad Sci U S A **2019**;116(52):26798-807 doi 10.1073/pnas.1915608116.

9.	Abu Rayyan A, Kamal L, Casadei S, Brownstein Z, Zahdeh F, Shahin H*, et al.* Genomic analysis of inherited hearing loss in the Palestinian population. Proc Natl Acad Sci U S A **2020**;117(33):20070-6 doi 10.1073/pnas.2009628117.

10.	Jaganathan K, Kyriazopoulou Panagiotopoulou S, McRae JF, Darbandi SF, Knowles D, Li YI*, et al.* Predicting Splicing from Primary Sequence with Deep Learning. Cell **2019**;176(3):535-48.e24 doi 10.1016/j.cell.2018.12.015.

11.	Wappenschmidt B, Becker AA, Hauke J, Weber U, Engert S, Köhler J*, et al.* Analysis of 30 putative BRCA1 splicing mutations in hereditary breast and ovarian cancer families identifies exonic splice site mutations that escape in silico prediction. PLoS One **2012**;7(12):e50800 doi 10.1371/journal.pone.0050800.
