## Supplementary Figures for "*BRCA1* secondary splice-site mutations drive exon-skipping and PARP inhibitor resistance"

Nesic et al., 2023

**Figure S1**

**A.**

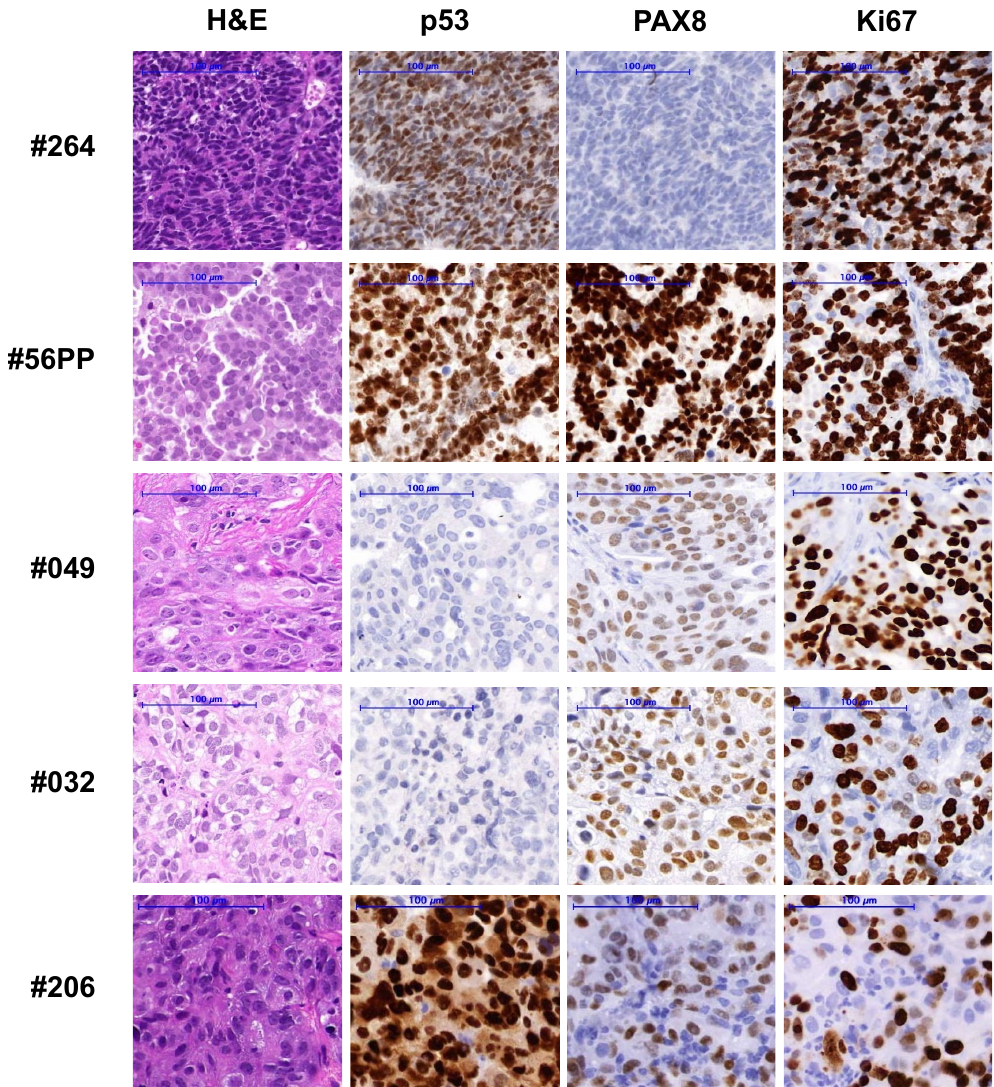

**B.**
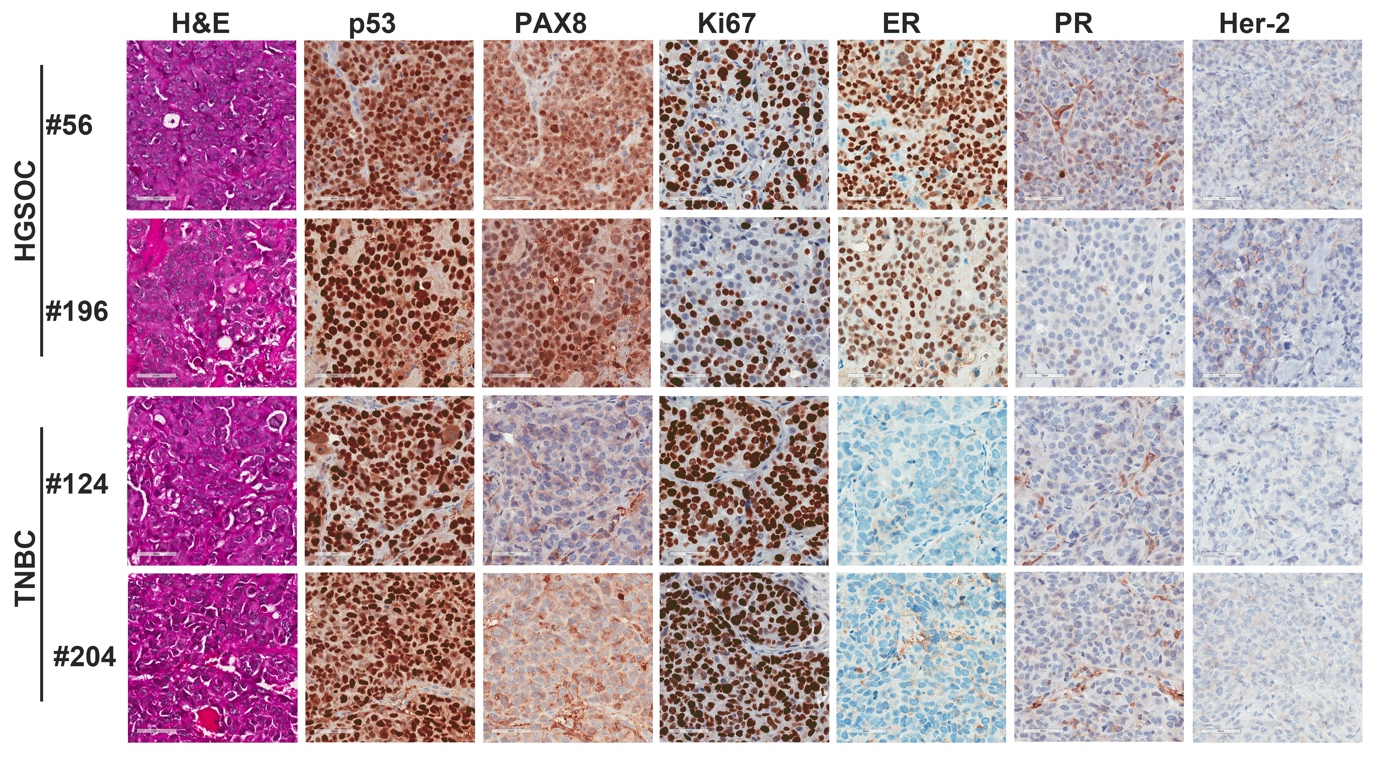

**Supplementary Figure S1. Histology of each PDX model.**

Immunohistochemistry (IHC) staining was used to assess PDX tumour morphology and histology. A. Histology of WEHI PDX. Hematoxylin and eosin (H&E) staining confirmed morphology consistent with HGSOC for models #56PP, #049, #032 and #206, and OCS for PDX model #264. PDX #56PP harbors a *TP53*:c.963delA (p.K321fs) mutation, but still had p53 staining possibly as this mutation occurs later in the protein and may not lead to nonsense-mediated decay. PDX #206 harbours a *TP53*:c.451C>T mutation (p.P151S) and had high p53 staining, while PDX #049 was found to have a frameshift *TP53*:c.428_429insCA (E144Sfs*27) mutation, leading to a pre-mature stop codon early in the protein and absent p53 staining. OCS PDX #264 had a *TP53*: c.1006G>T (p.E336X) mutation but still had p53 staining by IHC, possibly for the same reason as #56PP. High PAX8 was observed in HGSOC models #56PP, #049 #032 and #206, but none in OCS PDX model #264. Scale bars are 100μm. B. FCCC PDX models #56 and #196 were confirmed to have HGSOC histology, and PDX #124 and #204 were confirmed to have TNBC histology. Scale bar represents 50um.

**Figure S2**

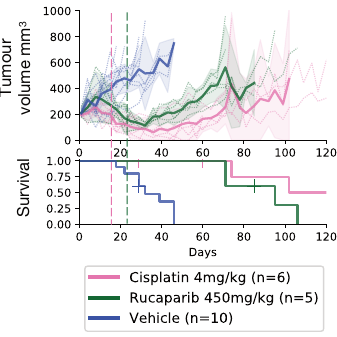

**Supplementary Figure S2. WEHI HGSOC PDX #56 lineage (#56C) treated with 4 mg/kg cisplatin remains sensitive to rucaparib and cisplatin.**

A new lineage of HGSOC PDX #56 (#56C) was derived from a mouse that had been treated with 4 mg/kg cisplatin, and this lineage was found to still be responsive to both rucaparib 450 mg/kg and cisplatin 4 mg/kg (P <0.001 for both compared with vehicle). Mean tumor volume (mm^3^) ± 95% CI (hashed lines are representing individual mice) and corresponding Kaplan–Meier survival analysis. Censored events are represented by crosses on Kaplan–Meier plot; n = individual mice.

**Figure S3**

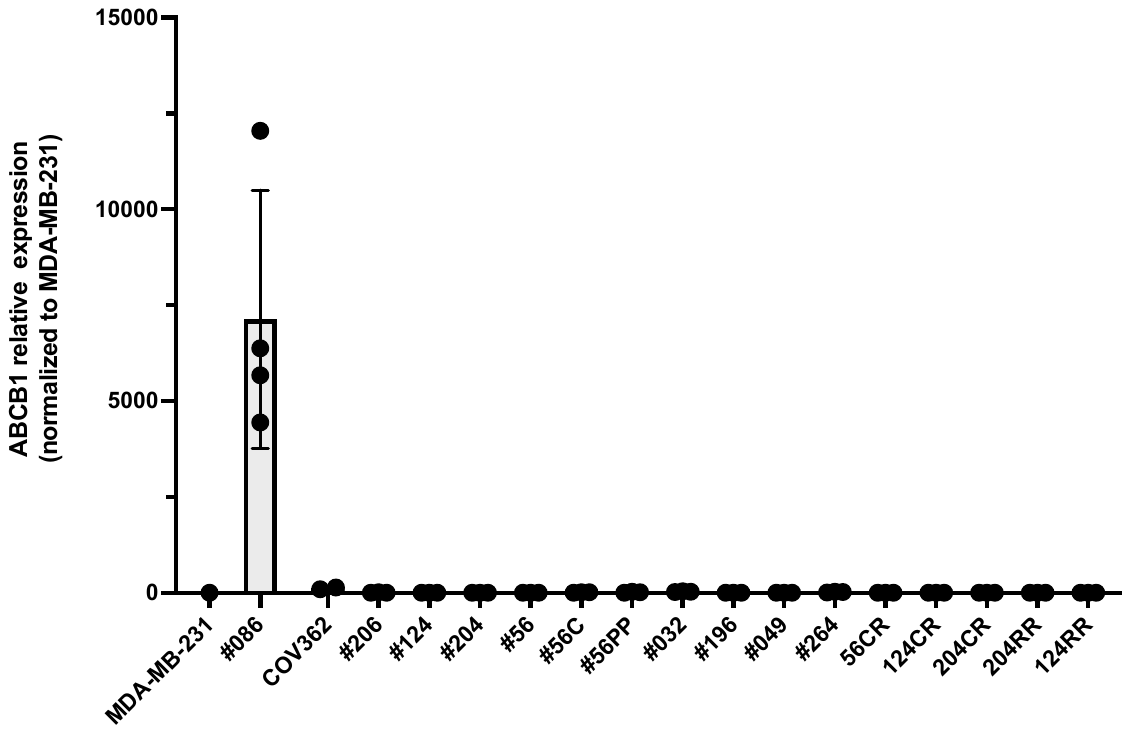

**Supplementary Figure S3. *ABCB1* gene expression in each PDX and cell line model.**

Expression of the drug efflux gene *ABCB1* was assessed by two-step RT-qPCR for each model in our study to exclude this as a mechanism of PARP inhibitor resistance. None of the models in our study had high *ABCB1* expression relative to the *ABCB1*-high control PDX #086. Data presented is fold-change relative to cell line MDA-MB-231. Each point represents an independent PDX or cell line passage, with mean ± standard deviation presented.

**Figure S4**

**A.**

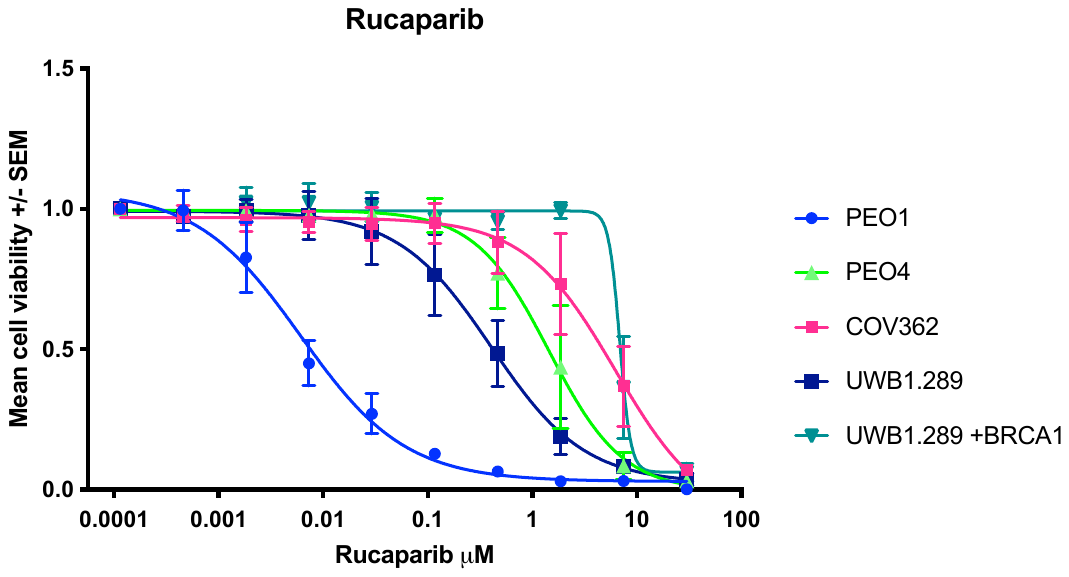

**B.**

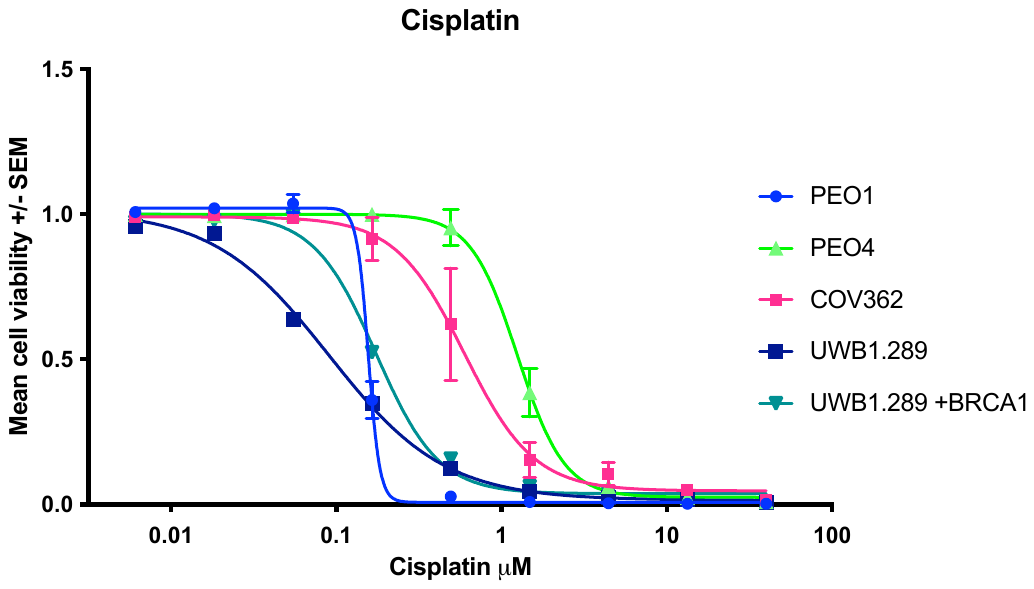

**Supplementary Figure S4. COV362 cell line is resistant to PARPi and platinum.**

The CTG cell viability assays demonstrated that Δ11q-high cell line COV362 is resistant to (A) PARPi rucaparib and (B) platinum treatment relative to the sensitive control cell line PEO1 (*BRCA2*-mutant), and resistant control cell lines PEO4 (*BRCA2* reversion mutation, HR competent), UWB1.289 (Δ11q-high) and UWB1.289+BRCA1 (UWB1.289 harbouring a *BRCA1* expression vector – additional resistance to PARPi compared to the parental line). Data presented is mean +/- SEM from n=3 independent experiments.

**Figure S5**

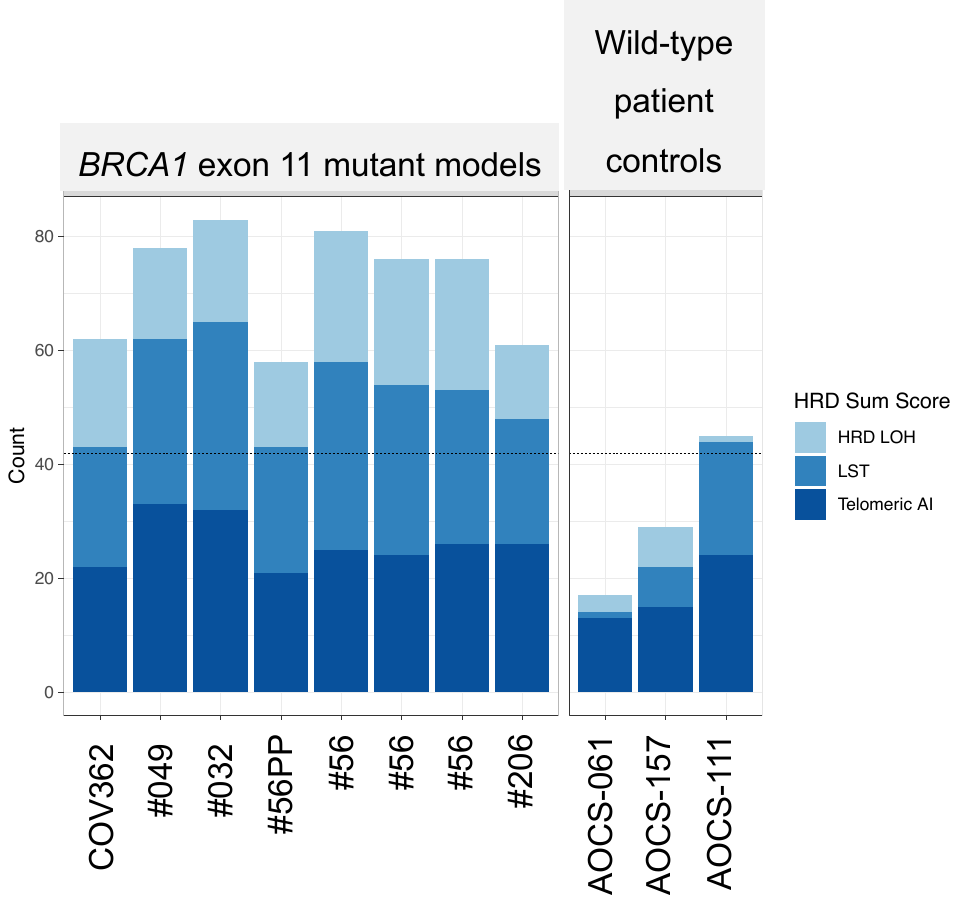

**Supplementary Figure S5. SNP array HRD scores for PDX and cell line COV362.**

All Δ11/Δ11q-high models (PDX and cell line COV362) that were tested SNP arrays in this study were found to have high combined HRD scores, indicating either current or historical HRD in the tumour. Dashed line indicates previously defined threshold (1-3). SNP arrays of AOCS were performed previously in the ICGC-Ovarian Cancer Project (4) and were re-analysed as controls for this study.

**Figure S6**

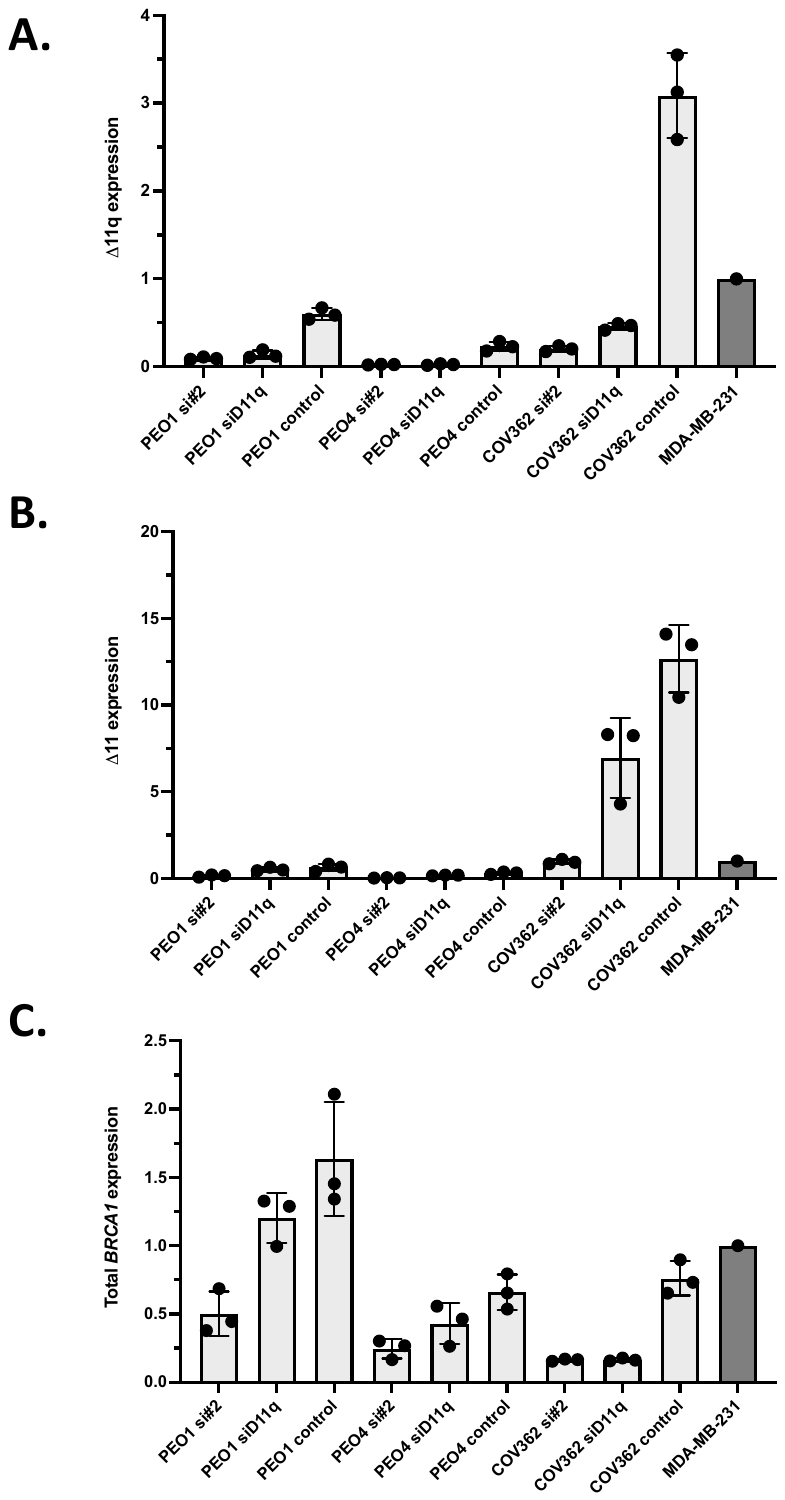

**Supplementary Figure S6. *BRCA1* isoform expression following siRNA treatment.**

Following siRNA treatment targeting either most *BRCA1* isoforms (si#2), Δ11q isoform specifically (siD11q) or a non-specific control, the expression of the (A) Δ11q isoform, (B) Δ11 isoform or (C) total *BRCA1* expression was assessed using two-step RT-qPCR. Data presented is fold-change relative to cell line MDA-MB-231. Each point represents an independent experiment, with mean ± standard deviation presented.

**Figure S7**

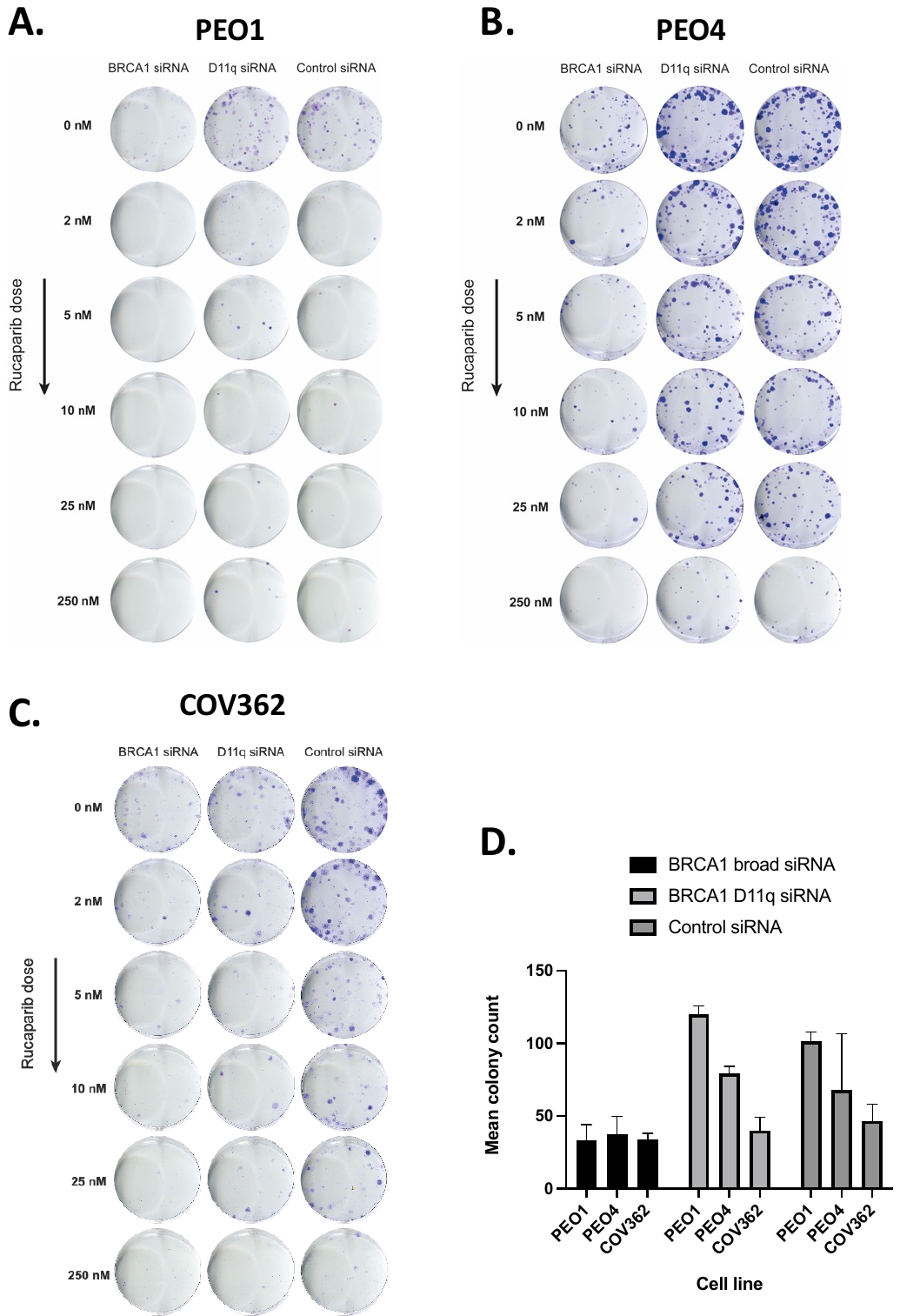

**Supplementary Figure S7. Colony formation of COV362 following siRNA and PARPi treatments.**

Representative images showing colony formation of (A) HR deficient control cell line PEO1, (B) HR competent control cell line PEO4 and (C) Δ11q-high cell line COV362 following siRNA knockdown of most BRCA1 isoforms (BRCA1 siRNA), Δ11q specifically or control siRNA treatment and various doses of rucaparib. (D) Mean colony count for rucaparib- untreated wells (± standard deviation) shows that broad *BRCA1* siRNA treatment alone reduces colony-forming capacity in PEO1 and PEO4.

**Figure S8**

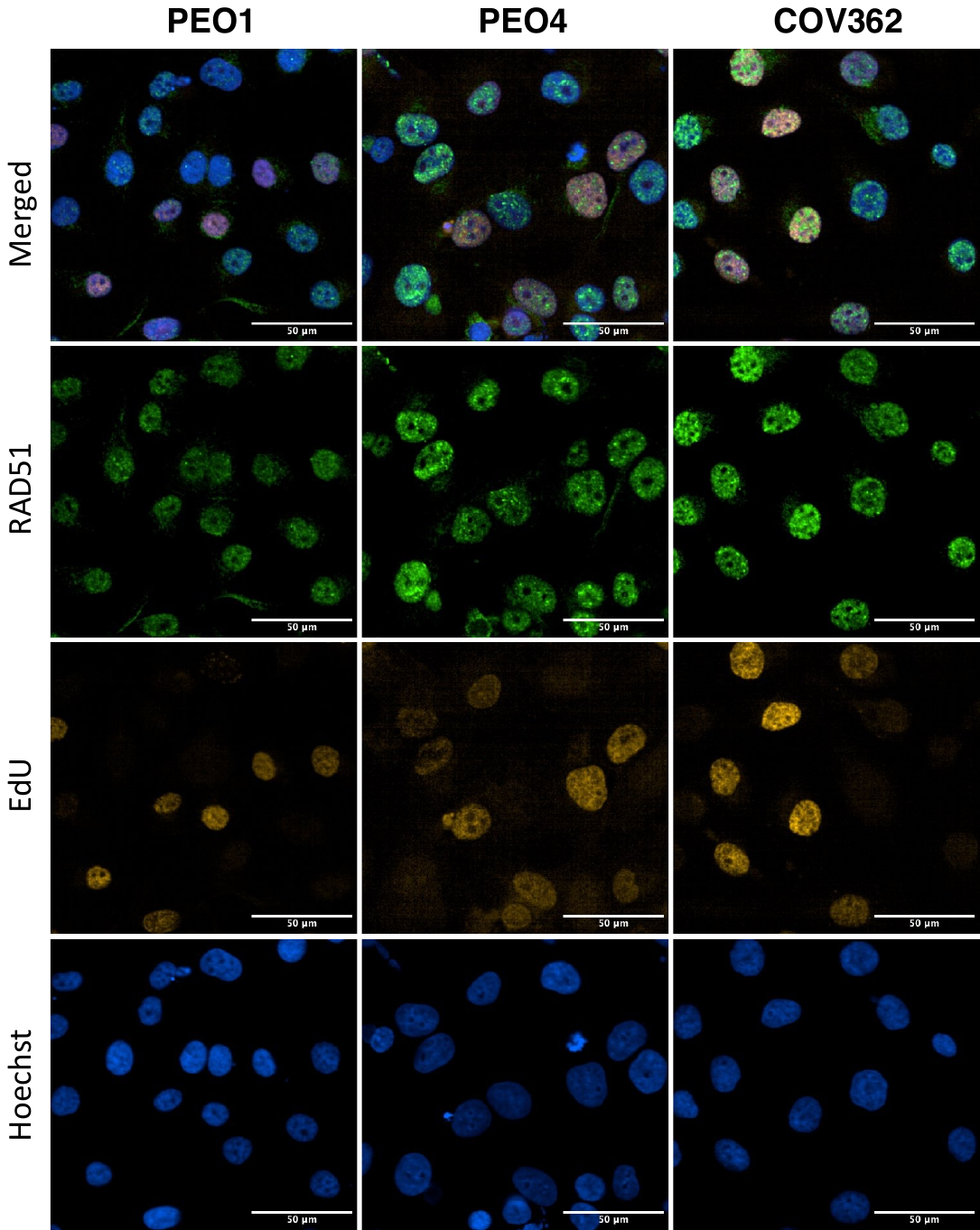

**Supplementary Figure S8. RAD51 foci formation in cell line COV362.**

Representative images showing RAD51 foci formation 1 hour post 10Gy γ-irradiation in cell lines COV362, HR deficient control PEO1 and HR competent control PEO4. Scale bar is 50μm.

**Figure S9**

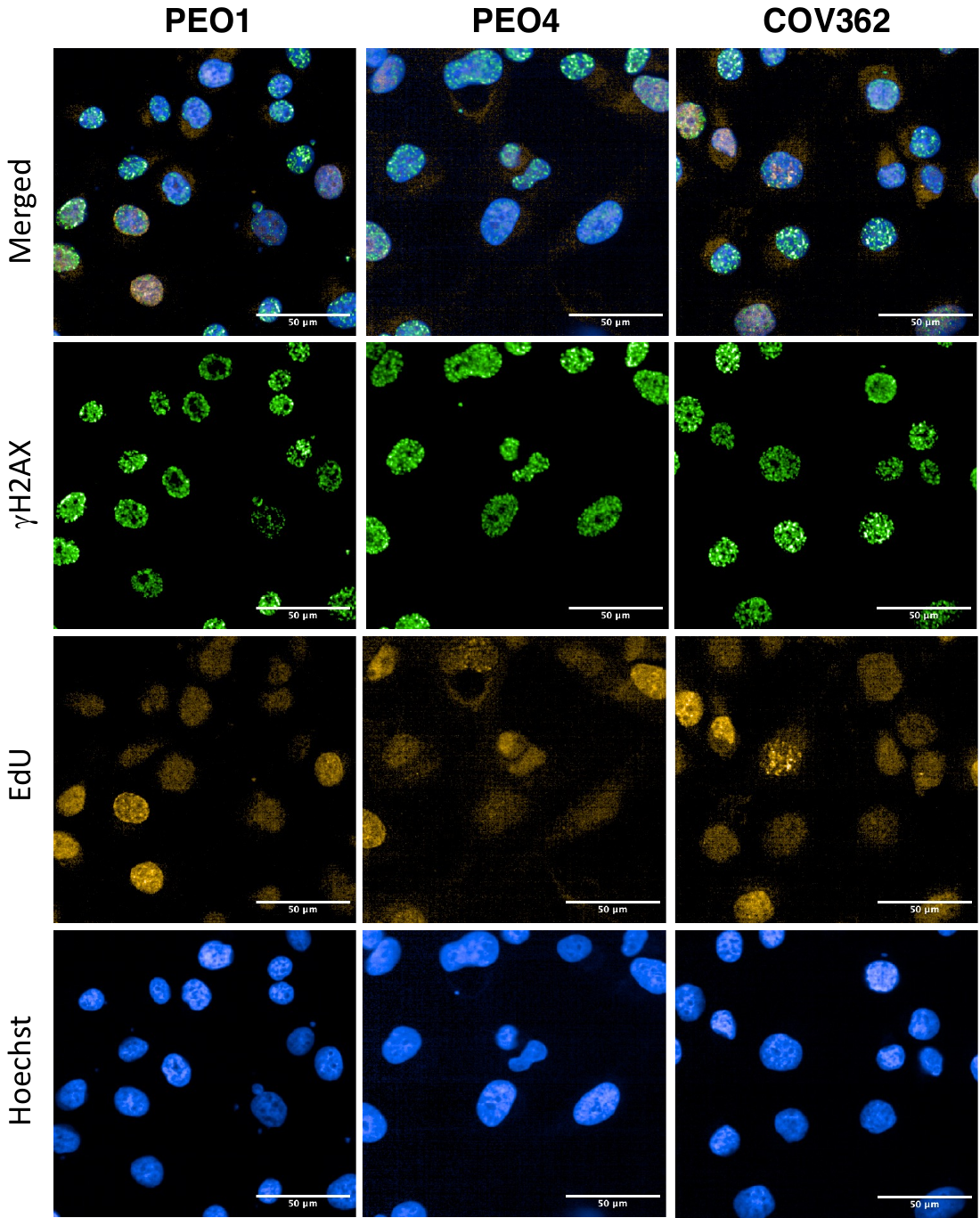

**Supplementary Figure S9. γH2AX foci formation in cell line COV362.**

Representative images showing γH2AX foci formation 1 hour post 10Gy γ-irradiation in cell lines COV362, HR deficient control PEO1 and HR competent control PEO4. Scale bar is 50μm.

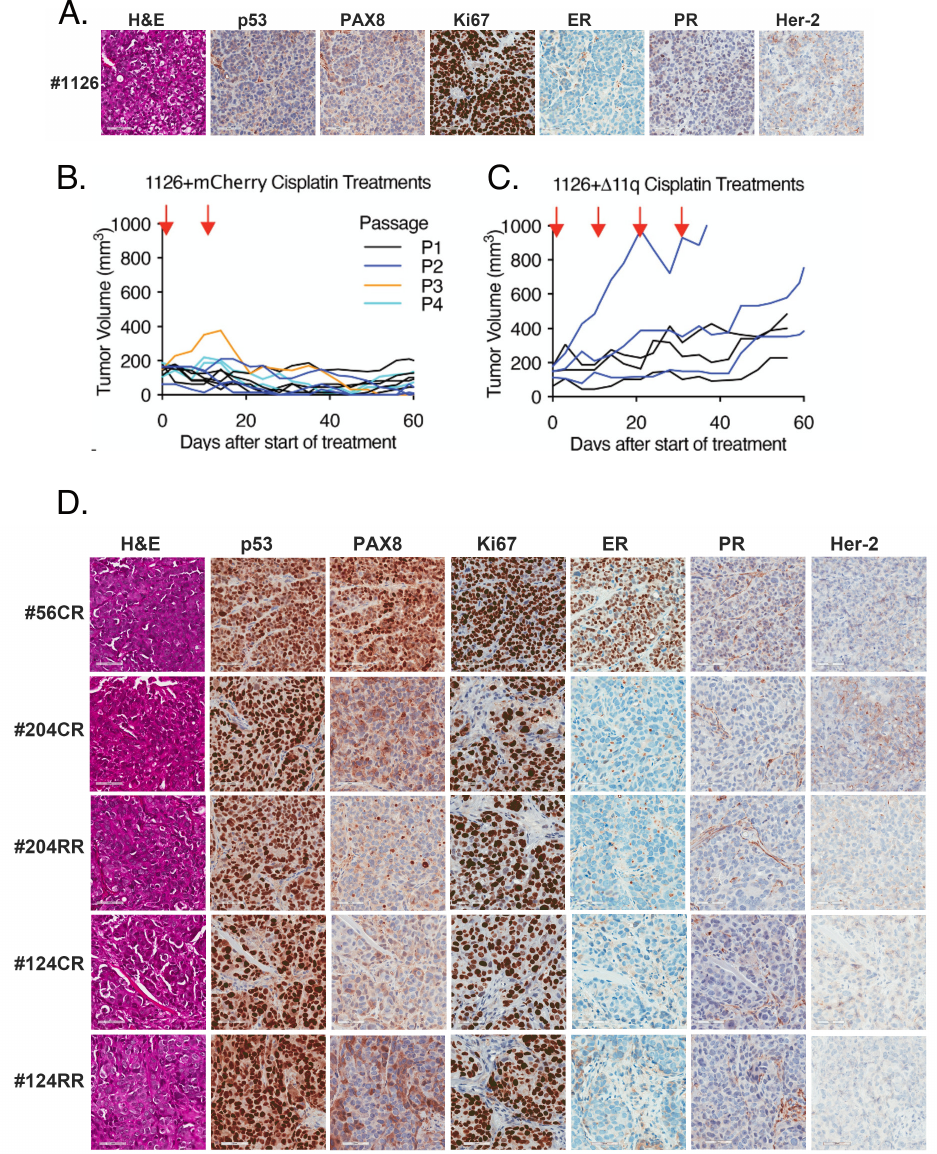

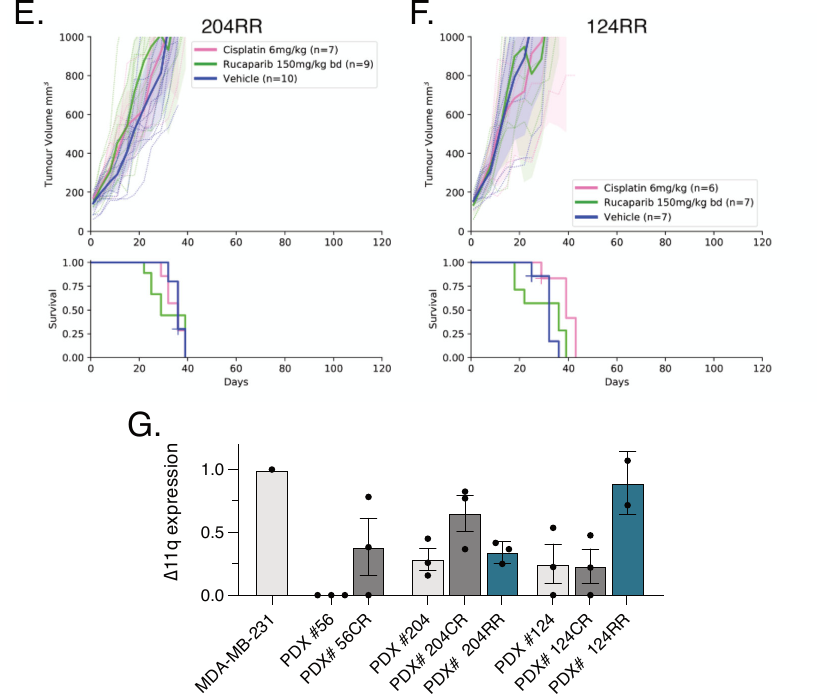

**Supplementary Figure S10. Generation of cisplatin and rucaparib resistant FCCC PDX lineages.**

(A) IHC confirmed that PDX #1126 has a TNBC histology. (B) PDX #1126 harbors a truncating exon 13 mutation in *BRCA1* and produces no detectable BRCA1 protein. PDX #1126 tumors were infected with lentivirus encoding an mCherry control and subjected to four passages of cisplatin treatments in mice as described in Figure 4H. Tumor volumes were measured 2-3 times per week and are presented for individual tumors with color indicating the tumor passage and cisplatin treatments shown by red arrow. After four passages in NSG mice, PDX #1126+mCherry tumors remained exquisitely sensitive to cisplatin and no resistant derivative emerged. (C) Cisplatin resistance was selected for in PDX #1126 tumors infected with lentivirus encoding BRCA1-Δ11q as described for the mCherry control in A. Unlike the mCherry control, resistant tumors emerged after the second passage for tumors expressing BRCA1-Δ11q. (D) PDX lineages #56CR, #204CR, #204RR, #124CR and #124RR all matched their corresponding parental PDX histology (Figure S1B). (E) Rucaparib resistant PDX #204 (PDX #204RR) were obtained after four passages of rucaparib treatments using the scheme described in Figure 4H. The resultant resistant derivative was assessed for response to rucaparib and cisplatin using a single dose of 6 mg/kg cisplatin or 150 mg/kg bid rucaparib for 10 days. Unlike parental PDX #204 tumors, PDX #204RR and PDX #204CR, tumors rapidly grew through both cisplatin and rucaparib treatments. (F) Rucaparib resistant PDX #124 (PDX #124RR) were obtained after six passages of rucaparib treatments. Similar to PDX #124CR, PDX #124RR lacked response to cisplatin or rucaparib while the parental PDX #124 was responsive to both therapies. Treatments were performed as described in C. (G) *BRCA1* Δ11q qPCR of the resulting CR and RR PDX lineages reveals increased Δ11q mRNA expression in #56CR, #204CR and #124RR, however these difference were not found to be statistically significant when compared to the parental lineage (Mean ± SEM shown; Welch two-tailed t test). Results for #56CR are concordant with western blot results (Figure 4O), but not so for other PDX lineages. This may be due to differences in mRNA processing between lineages, or due to tumour heterogeneity.

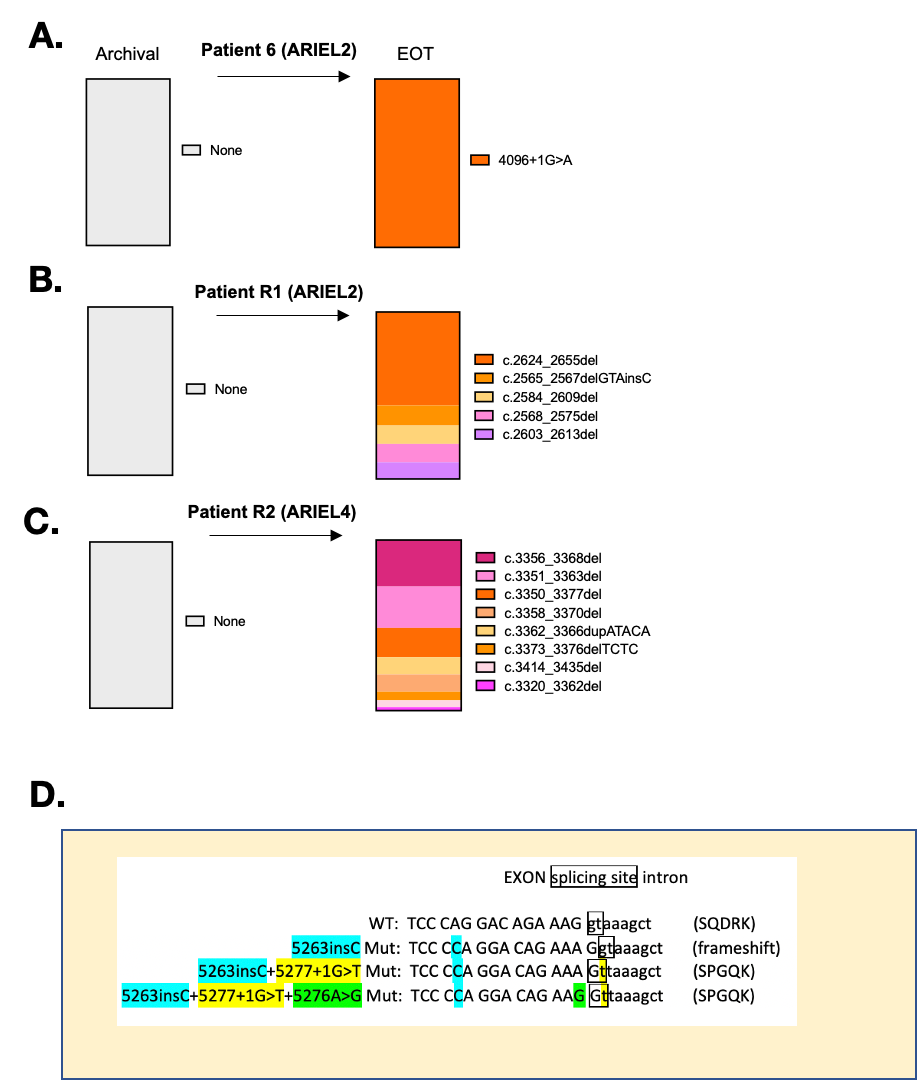

**Supplementary Figure S11. Interpreting *BRCA1* secondary mutations found in ARIEL2 and ARIEL4 clinical trial patient samples.**

(A) Patient 6 was platinum sensitive (2 prior lines of platinum) and had a partial response to rucaparib on the ARIEL2 clinical trial. The *BRCA1* exon 11 splice site mutation 4096+1G>A was detected in post-treatment tissue sample by FMI, but not in an archival sample. Deleterious germline mutation was c.2042_2043insT, in exon 11. (B) and (C) show examples of patients that had multiple non-splicing secondary mutations arise in their plasma samples following rucaparib treatment, showing that this number of reversion events is not unique to patients with secondary *BRCA1* splice-site mutations (*e.g.* patients 3 and 4). (D) The *BRCA1* exon 20 c.5276A>G mutation (in green) was found to co-occur in all cases with a primary c.5266dupC mutation (in blue) and a c.5277+1G>T secondary mutation (in yellow). c.5276A>G was confirmed not to be an artefact, thus it is possible that c.5277+1G>T generates a new GT splice site that is shifted 1 bp from the original position into the exon, effectively creating an additional frameshift (exon loses 1 bp, intron gains 1 bp) and restoring the reading frame. The 5276A>G mutation may be required to re-establish the boundary sequence adjacent to the splice site to enable proper splicing.

**References**

1.	Mills GB, Timms KM, Reid JE, Gutin AS, Krivak TC, Hennessy B*, et al.* Homologous recombination deficiency score shows superior association with outcome compared with its individual score components in platinum-treated serous ovarian cancer. Gynecologic Oncology **2016**;141:2-3 doi 10.1016/j.ygyno.2016.04.034.

2.	Telli ML, Timms KM, Reid J, Hennessy B, Mills GB, Jensen KC*, et al.* Homologous Recombination Deficiency (HRD) Score Predicts Response to Platinum-Containing Neoadjuvant Chemotherapy in Patients with Triple-Negative Breast Cancer. Clin Cancer Res **2016**;22(15):3764-73 doi 10.1158/1078-0432.CCR-15-2477.

3.	Sztupinszki Z, Diossy M, Krzystanek M, Reiniger L, Csabai I, Favero F*, et al.* Migrating the SNP array-based homologous recombination deficiency measures to next generation sequencing data of breast cancer. npj Breast Cancer **2018**;4(1):16 doi 10.1038/s41523-018-0066-6.

4.	Patch AM, Christie EL, Etemadmoghadam D, Garsed DW, George J, Fereday S*, et al.* Whole-genome characterization of chemoresistant ovarian cancer. Nature **2015**;521(7553):489-94 doi 10.1038/nature14410.
